# Clinical validation of a novel metagenomic nanopore sequencing method for detecting viral respiratory pathogens: diagnostic accuracy study

**DOI:** 10.64898/2026.02.06.26345651

**Authors:** Nicholas D. Sanderson, Kate E. Dingle, Katie M.V. Hopkins, Ali Vaughan, Matthew Colpus, Melody Parker, Elisabeth Vilde Dietz, Jessica Gentry, Anita Justice, Sarah Oakley, Lucinda Barrett, T. Phuong Quan, Nicole Stoesser, David W. Eyre, Philip Bejon, Ann Sarah Walker, Bernadette C. Young

## Abstract

**Background:** Clinical metagenomics (CMg) offers high-throughput respiratory pathogen detection with a wider range than targeted, probe-dependent diagnostics. Sequencing cost and the challenges of high host biomass in non-invasive samples are barriers to the use of CMg in high-throughput respiratory pathogen detection.

**Methods:** We optimised a long-read sequencing workflow to detect RNA viruses in nasopharyngeal swabs, employing pathogen enrichment and ONT sequencing. As a pre-requisite for agnostic pathogen detection, we first derived quality control (QC) criteria and diagnostic thresholds against a gold-standard comprising 23 pathogen targets detected by routine multiplex PCR. We validated this workflow using 344 prospectively collected upper respiratory tract samples submitted for routine testing.

**Findings:** Using pre-defined QC and positivity criteria, the workflow’s sensitivity versus PCR was 51% (95%CI: 45%-57%) (133/260 positive targets detected) (ranging from 19%-85% across pathogens with >20 gold-standard detections), and specificity 99.8% (95%CI: 99.6%-99.9%) (3836/3845 negative targets not detected). Sensitivity improved to 58% using post-hoc optimised thresholds, 61% only considering RNA pathogens, 70% excluding rhinovirus/enterovirus and 83% excluding samples with qPCR Ct values ≥35. Read crossover from multiplex sequencing contributed most (7/9) false-positives: only 2 plausible additional pathogens were identified (rhinovirus and coronavirus OC43). 41 respiratory syncytial virus (RSV), 13 influenza A and 10 rhinovirus/enterovirus were successfully sub-typed by sequencing. Multiplexed nanopore sequencing costs were £112/sample.

**Interpretation:** Although CMg has substantial diagnostic potential, this validation study demonstrates the technical limitations of current metagenomic sequencing methods applied to viral detection in upper respiratory tract samples with high host and low pathogen biomass. Its greater sensitivity at higher viral loads demonstrates the importance of identifying the most appropriate use cases to maximise its utility and value.

**Funding:** National Institute for Health and Care Research Oxford Biomedical Research Centre.

**Research in context:** *Evidence before this study:* Clinical metagenomics (CMg) provides the potential for rapid respiratory pathogen detection with a wider range than targeted, probe-dependent diagnostics. Searching PubMed for studies published between Jan 1 2018 and December 1 2025 using the terms ‘respiratory tract infection’ AND ‘metagenomics’ AND ‘diagnosis’, with no language restrictions yielded 734 items. We identified 15 clinical studies assessing the diagnostic performance of a metagenomic workflow for respiratory samples. Only 3 of these included upper respiratory tract samples (nasopharyngeal swabs); the remaining studies exclusively investigated invasive samples from the lower respiratory tract (bronchioalveolar lavage fluid). One of the three relevant studies assessed detection of viral pathogens in 83 specimens from critical care patients as part of pan-microbial nanopore sequencing metagenomic assay; only 2 samples included were nasopharyngeal samples. The second validated a high-depth short-read CMg workflow in 191 samples (85% nasopharyngeal), reporting 93.6% sensitivity and 98.8% specificity across eight respiratory pathogens detected by respiratory PCR. This workflow required substantial equipment and sequencing costs. A third study validated a nanopore sequencing CMg workflow, with lower laboratory footprint and sequencing costs than the second study. In 359 nasopharyngeal samples, they reported 61% sensitivity and 100% specificity against four respiratory viral targets detected by PCR. The costs of CMg mean that routine deployment as a high-throughput test with broad use will require demonstrating good performance across a range of common respiratory pathogens (such as can be assayed with commercially available extended multiplex PCR testing) and in upper respiratory tract samples.

*Added value of this study:* This prospective study is the first to evaluate nanopore sequencing for detecting a broad range of common respiratory viruses in nasopharyngeal samples. We validated a CMg workflow employing a modernised SISPA step for viral amplification, in 344 prospectively collected non-invasive samples, from which 19 different respiratory pathogens were detected by routine PCR testing. Using pre-defined bioinformatic thresholds overall sensitivity was 51% compared with PCR. This could be improved to 83% by limiting analysis to RNA viruses with Ct <35 and using post-hoc exploratory bioinformatic criteria. Sequencing costs were 45% lower than short-read sequencing.

*Implications of all the available evidence:* Our results demonstrate rigorous bioinformatic thresholds are essential in multiplexed CMg sequencing to reduce false-positive detections, a particular danger with imperfect barcode de-multiplexing. However these thresholds impair true-positive detection in samples with a high ratio of host-to-pathogen biomass. Further research should focus on identifying in which samples and clinical settings CMg can offer greater value in patient care.

## Introduction

Respiratory tract infections remain a leading cause of illness with substantial impact on healthcare systems.^1^ Diagnostic testing for respiratory infections continues to pose major challenges.^2,3^ Much routine testing remains targeted,^4^ testing for either a single pathogen, or configured as ‘syndromic’ panels testing pre-defined pathogen arrays.^5,6^ While sensitive, these assays are constrained in scope,^7^ are vulnerable to naturally occurring variations in the assay target,^8,9^ and may fail to distinguish between important species or subtypes.

Clinical metagenomics (CMg) has emerged as a promising modality for rapid, untargeted and simultaneous pathogen detection directly from clinical samples. Studies reporting CMg use in critical care have focussed on lower respiratory tract specimens such as bronchoalveolar lavage (BAL),^10,11^ reporting performance for bacterial detection and antimicrobial resistance profiling.^12,13^ However, use can extend to viral and fungal infections.^14^

Technical challenges limit broader routine application of respiratory CMg. Non-invasive naso-pharyngeal swabs contain high ratios of host to pathogen biomass, making accurate detection of signal from noise problematic: some CMg workflows count only a single pathogen read as detection.^15,16^ Dominant human nucleic acid requires depletion strategies to increase relative pathogen abundance.^16^ Short-read sequencing to very high depths can overcome these challenges, but at a high price: sequencing costs of a well-validated workflow were $260USD/test (estimated £198GBP on 2025 average exchange rate).^17^ Pathogen enrichment strategies include targeted baits, or untargeted amplification (sequence independent, single primer amplification (SISPA), or phi29 polymerase).^18,19^ Further, contaminating material and barcode/index misclassification can all produce false-positive results,^20^ requiring optimised laboratory practices and bioinformatic processes.^21^

Whilst ultimately the CMg goal is agnostic pathogen detection, validation of sensitivity and specificity against known pathogen targets is necessary (but not sufficient) to assess performance. We therefore undertook a clinical evaluation of an end-to-end workflow for detecting common RNA respiratory viruses from nasopharyngeal samples. The workflow combines lower-cost virus enrichment and non-target depletion approaches, and Oxford Nanopore Technologies (ONT) sequencing. We used a derivation sample set to develop bioinformatic thresholds, and then evaluated their performance in an independent validation sample set, using commercial multiplex PCR assays as the comparator to quantify sensitivity and specificity, and employing multiple laboratory and bioinformatic quality controls.

## Methods

### Respiratory samples for pathogen detection

Nasopharyngeal samples included in this study were submitted in viral transport medium for testing in the Microbiology Laboratory, Oxford University Hospitals NHS Foundation Trust (Oxford, UK). PCR was performed with three platforms: two four-target assays (Alinity Resp-4-plex assay (Abbott, USA), or Xpert Xpress CoV-2/Flu/RSV plus (Cepheid, USA)), or a 23-target assay (BIOFIRE FilmArray Respiratory 2.1 panel (Biomerieux, France)) (**Tab.S1**), following manufacturer’s instructions. The four-target assays (covering influenza A/B, respiratory syncytial virus (RSV), and severe acute respiratory syndrome coronavirus 2 (SARS-CoV-2)) were performed routinely, while 23-target PCR testing was undertaken for samples from immunocompromised patients or those admitted to critical care. Samples surplus to routine testing were retrieved. Inclusion criteria were: (i) nose and throat swabs, (ii) <7 days since patient sampling, iii) pathogen detected at cycle threshold (Ct) or cycle number (Cn) value ≤30 for Resp-4-Plex and Xpert assays or sampled tested by BIOFIRE FilmArray Respiratory 2.1 Panel (no Ct/Cn available). For approximately every eight positive samples, one FilmArray negative sample was collected (**Tab.1(a))**.

**Table 1.** Characteristics of patient samples (nose and throat swabs) and sequences in derivation and validation sets.

**(a)**
|  | Derivation set samples (N=38) | Derivation set sequences† (N=104) | Derivation set sequences QC pass†† (N=61) | Validation set samples/ sequences (N=344) | Validation set samples/ sequences QC pass†† (N=273) |
| --- | --- | --- | --- | --- | --- |
| Original test by (n (%)) |  |  |  |  |  |
| Xpert Xpress CoV-2/Flu/RSV plus* | 2 (5%) | 4 (4%) | 4 (7%) | 9 (3%) (2-°,7+) | 9 (3%) (2-°,7+) |
| Alinity Resp-4-Plex* | 6‡ (16%) | 18‡ (17%) | 8‡ (13%) | 136 (40%) (0-,136+) | 102 (37%) (0-,102+) |
| BIOFIRE FilmArray Respiratory 2.1** | 31‡ (82%) | 85‡ (82%) | 51‡ (84%) | 199 (58%) (45-,154+) | 162 (59%) (36-,126+) |
| Number of pathogen targets detected‡‡, n (%) |  |  |  |  |  |
| 0 | 0 | 0 | 0 | 47 (14%) | 38 (14%) |
| 1 | 34 (89%) | 94 (90%) | 55 (90%) | 270 (78%) | 212 (78%) |
| 2 | 4 (11%) | 10 (10%) | 6 (10%) | 25 (7%) | 21 (8%) |
| 3 | 0 | 0 | 0 | 2 (1%) | 2 (1%) |
| Total pathogen targets detected | 42 | 114 | 67 | 326 | 260 |
| Thousand reads generated | - | 92 (69-127) | 89 (68-108) | 115 (81-151) | 115 (82-146) |
| Median read length | - | 292 (278-308) | 290 (275-300) | 270 (246-292) | 276 (256-295) |
| N50 | - | 428 (375-469) | 448 (382-473) | 386 (336-429) | 393 (348-432) |
| Read PHRED quality | - | 10.9 (10.0-12.3) | 10.5 (9.9-11.2) | 11.4 (11.0-11.7) | 11.4 (11.1-11.7) |
| Any target pathogen reads identified, n (%) | - | 77 (74%) | 49 (80%) | 230 (67%) | 191 (70%) |
| - median (IQR) reads if >0 | - | 35 (5-166) | 18 (5-98) | 6 (2-151) | 8 (2-188) |
| Any gold standard (target) pathogen reads identified, n (%) | - | 64 (62%) | 45 (74%) | 198 (58%) | 162 (59%) |
| - median (IQR) reads if >0 | - | 70 (6-2817) | 46 (6-135) | 44 (4-514) | 58 (4-603) |

\* tests for influenza A/B, RSV and SARS-CoV-2 (**Tab.S1**).
\*\* tests for adenovirus, coronaviruses (229E, HKU1, NL63, OC43, SARS-CoV-2, MERS-CoV), human metapneumovirus, human rhino/enterovirus, influenza viruses (including subtypes influenza A, A/H1, A/H3, A/H1-2009), influenza B virus, parainfluenza viruses (1-4), respiratory syncytial virus, *Bordetella parapertussis*, *Bordetella pertussis*, *Chlamydia pneumoniae* and *Mycoplasma pneumoniae* (**Tab.S1**).
† Original derivation samples were split and sequenced multiple times. Some sequenced samples included pooled samples tested with multiple platforms, see ‡. For the validation set each sample was sequenced once only.
†† Samples that passed run, batch and sample internal controls, see **Tab.2**.
‡ One derivation set sample was pooled and tested using both Resp-4-plex and FilmArray; this was sequenced 3 times.

|  | Validation set samples/<br>sequences (N=344) | Validation set samples/<br>sequences QC pass†† (N=273) |
| --- | --- | --- |
| Patient age (years) | 48 (3-70) n=342* | 38 (2-68) n=271* |
| <1 | 46 (13%) | 37 (14%) |
| 1-12 | 70 (20%) | 65 (24%) |
| 13-49 | 56 (16%) | 45 (17%) |
| 50-70 | 86 (25%) | 66 (24%) |
| 71-84 | 58 (17%) | 39 (14%) |
| 85+ | 26 (8%) | 19 (7%) |
| Male | 176 (51%) n=342* | 141 (52%) n=271* |
| Location where sample taken |  |  |
| Haematology or oncology | 103 (30%) | 83 (30%) |
| Other inpatient adult ward | 77 (22%) | 53 (19%) |
| Other inpatient paediatric ward | 38 (11%) | 30 (11%) |
| Ambulatory or emergency department | 92 (27%) | 76 (28%) |
| Critical care | 19 (6%) | 18 (7%) |
| Community | 13 (4%) | 11 (4%) |
| Trial | 2 (1%) | 2 (1%) |
| Days between collection and extraction date | 5 (3-6) | 5 (3-6) |
| Copy number (Cn) (Alinity) | 22.5 (19.7-25.3) | 22.9 (20.3-25.5) |
| Cycle threshold (Ct) (Xpert) | 19.5 (18.8-22.1) | 19.5 (18.8-22.1) |
| Total pathogens detected by routine testing | 326 | 260 |
| Study qPCR performed** | 306 (94%) | 244 (94%) |
| Study qPCR run, but Ct undetermined (>40)<br>(% of routine testing targets with qPCR) | 68 (22%) | 50 (20%) |
| Study qPCR run, Ct determined (<40) (% of<br>routine testing targets with qPCR)<br>- Median (IQR) Ct | 238 (78%)<br>30.2 (IQR: 27.2-33.7) | 194 (80%)<br>30.0 (IQR: 27.2-33.9) |

\* missing values from anonymised trial samples. Percentages of non-missing values.
\*\* DNA target or multiple pathogens identified on routine testing and insufficient sample to run qPCR for all (see **supplemental methods**)
Note: showing n (%) or median (IQR). QC=quality control. See **Fig.S1** for run characteristics.

### Procedures

A derivation set was used to define bioinformatic criteria for pathogen detection by sequencing and thresholds for quality control (QC) for both positive and negative controls, criteria which were then fixed and tested using the validation set, with all validation assays and sequencing conducted by laboratory staff blinded to validation study results throughout. Validation sample size was determined by the duration of the winter 24/25 season and available staff.

Sample preparation and sequencing followed a single optimised laboratory method in the validation set, while derivation sample preparation and sequencing used a similar near final method (**Fig.1, supplemental methods**). Briefly, this comprised host depletion by low-speed centrifugation and nuclease treatment with M-SAN HQ (ArcticZymes), followed by viral precipitation with Intact Virus Precipitation Reagent (Thermo Fisher Scientific). Viral material was retrieved by high-speed centrifugation. RNA was extracted using Direct-zol RNA Microprep kit (Zymo Research), and underwent cDNA synthesis and SISPA. DNA libraries were prepared using Rapid Barcoding Kit and sequenced on R10.4.1 flow cells on GridION Mk1 (ONT).

**Figure 1.**
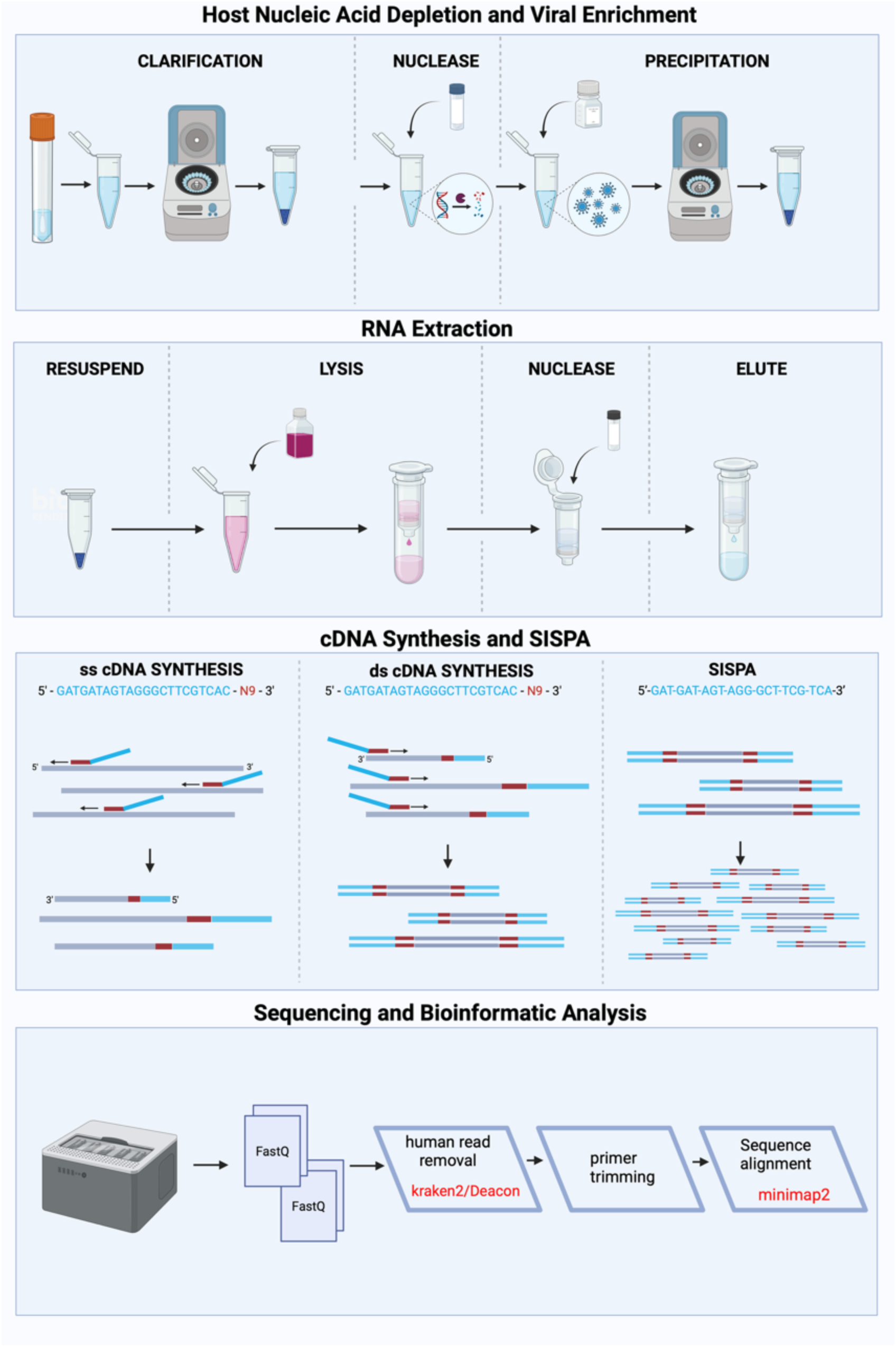
Clinical metagenomic workflow overview.

Samples were sequenced in batches with batch positive and negative controls, and internal amplification controls in each sample (**Tab.2**). Samples within both the derivation and validation sets were randomly mixed across sequencing runs. MS2 was added to each sample as a control of nucleic acid extraction and subsequent cDNA synthesis and SISPA. Three viruses were added to each sample following nucleic acid extraction as internal controls (ICs) of cDNA synthesis and SISPA. Each batch of 16 samples was accompanied by a positive amplification control consisting of the same 3 spiked-in viruses in a negative (no template) control. During cDNA synthesis, 3µl of single-stranded (ss) cDNA was reserved for quantification by qPCR (TaqMan Microbe Detection Assay, Thermo Fisher Scientific) targeting the respiratory virus(es) detected on gold-standard testing

**Table 2.** Quality check, controls and pathogen detection thresholds for CMg workflow.

| Type | Threshold for failure (any) | Applied to | Rationale |
| --- | --- | --- | --- |
| **Control (original prespecified before validation study)** |  |  |  |
| Run sequencing | <400 Mb data | Sequencing run (two batches of 16 samples plus controls) | Minimum level of sequencing data acquisition given variability across runs (**Figure S1**) |
| Sample sequencing | <25,000 reads | Each sample (unique barcode) | Minimum level of sequencing data acquisition given variability across samples. Minimum read number ensures sample QC is not biased by few very long reads (**Figure S1**) |
| Batch PCR negative control | $\geq 2$ reads mapped to any of the four positive spikes* or any target pathogen not failing (i.e. passing) main or ratio positivity criteria below | Negative control on each batch | Negative (no template) control included with all samples undergoing SISPA PCR in a batch. Detecting target amplification in this control indicates cross contamination. |
| Batch positive amplification control | <2 reads mapped to 2 or 3 of the 3 amplification controls | Positive control on each batch | Control with only 3 RNA virus spikes included in double stranded (ds) cDNA synthesis and SISPA amplification process for a batch of samples. Failure to detect the virus spike indicates these processes have failed across the batch. |
| Batch positive amplification control | MS2 or any target pathogen not failing (i.e. passing) main or ratio positivity criteria below | Positive control on each batch | Control with only 3 RNA virus spikes included in ds cDNA synthesis and SISPA amplification process for a batch of samples. Detecting MS2 or a target pathogen indicates cross-contamination. |
| Internal extraction, cDNA synthesis and SISPA control | <2 reads mapped to MS2** | Each sample | MS2 is added prior to RNA extraction. Failure to detect in a sample indicates that RNA extraction and/or subsequent amplification has failed within the sample |
| Internal cDNA synthesis and SISPA control | <2 reads mapped to 2 or 3 of the 3 amplification controls | Each sample | Three RNA virus spikes added prior to ds cDNA synthesis and downstream amplification. Failure to detect indicates these processes have failed within the sample. |
| **Positivity to a target pathogen (original pre-specified before validation study)** |  |  |  |
| Barcode crossover | <2 reads mapped to a target pathogen | Each sample | Reduce impact of barcode crossover, **Figure S2** |
| Main criteria | Area under the genome truncated at 10<br><0.003 AND coverage at depth 1 <0.25%<br>AND sample reads as a percentage of all<br>references <0.006% | Each sample | Combination of three metrics performance with high sensitivity and AUROC in derivation set, reducing potential impact of kitome and sample contamination observed in previous multiplexed sequencing studies. |
| Ratio criteria | Ratio sample reads as a percentage of all<br>references / area under the genome<br>truncated at 10<0.1 | Each sample | Excluded 23 false-positives in the full derivation set (**Figure 3**) |
| **Positivity to a target pathogen (alternative exploratory after validation study)** |  |  |  |
| Barcode crossover | <2 reads mapped to a target pathogen | Each sample | Reduce impact of barcode crossover, **Figure S2** |
| Main criteria | Sample reads as a percentage of reads<br>mapping to all target references <0.037% | Each sample | Single metric optimising sensitivity, specificity and AUROC and sensitivity in derivation set |

\* MS2 and internal viral amplification controls (Zika virus, murine respirovirus and orthoreovirus). Each sequencing run included 32 samples on a single flow cell. Samples were extracted and amplified in batches of 16 samples plus batch controls, and two batches included in a run.
\*\* Quality check included for the validation set only due to inconsistent performance of the MS2 stock in the derivation experiments.
Note: All run, batch individual sample quality check criteria had to pass in order to have pathogen reported as detected by sequencing; target pathogen declared present if all positivity criteria met. See **Fig.4** for a summary of the combinations of criteria passed and failed.

As the necessary first step, we targeted identification of 23 known pathogens/pathogen subtypes comprising the diagnostic targets of the commercial multiplex PCR platform (**Tab.S1**). To test the ability of the CMg workflow to reproduce PCR results at the highest possible resolution,^22^ pathogen detection was based on mapping reads to a curated database of matching known respiratory pathogen genome sequences (**Tab.S2**) using minimap2. Although the CMg workflow was focussed on RNA virus detection, we evaluated performance with respect to all FilmArray panel targets. We evaluated 28 bioinformatic metrics against true-positives and true-negatives to optimise detection (**Tab.S3**;**S4**), individually and in combination. Because each sample had a positive or negative result from PCR gold-standard testing for multiple pathogens which were or were not be detected by metagenomic sequencing, each sample contributed multiple observations to the analysis (i.e. each pathogen tested for using gold-standard PCR contributed one observation with the outcome being presence or absence of that pathogen on sequencing results). The analysis workflow is provided on https://github.com/oxfordmmm/AD_analysis.

## Results

In total 382 respiratory samples were retrieved, divided temporally into separate sample sets: a) a derivation (n=38 clinical samples, tested repeatedly to produce n=104 individual sequences); and b) a validation set (n=344 clinical samples, each tested once to produce n=344 individual sequences).

### Derivation Set: Defining thresholds for quality control

The derivation set included 104 sequencing results from 38 samples, all with pathogens detected on multiplex PCR testing (**Tab.1**, **Fig.2**). Total data obtained per run and reads per sample (**Fig.S1**) were used to set heuristic thresholds for sequencing run quality checks (**Tab.2**).

**Figure 2.**
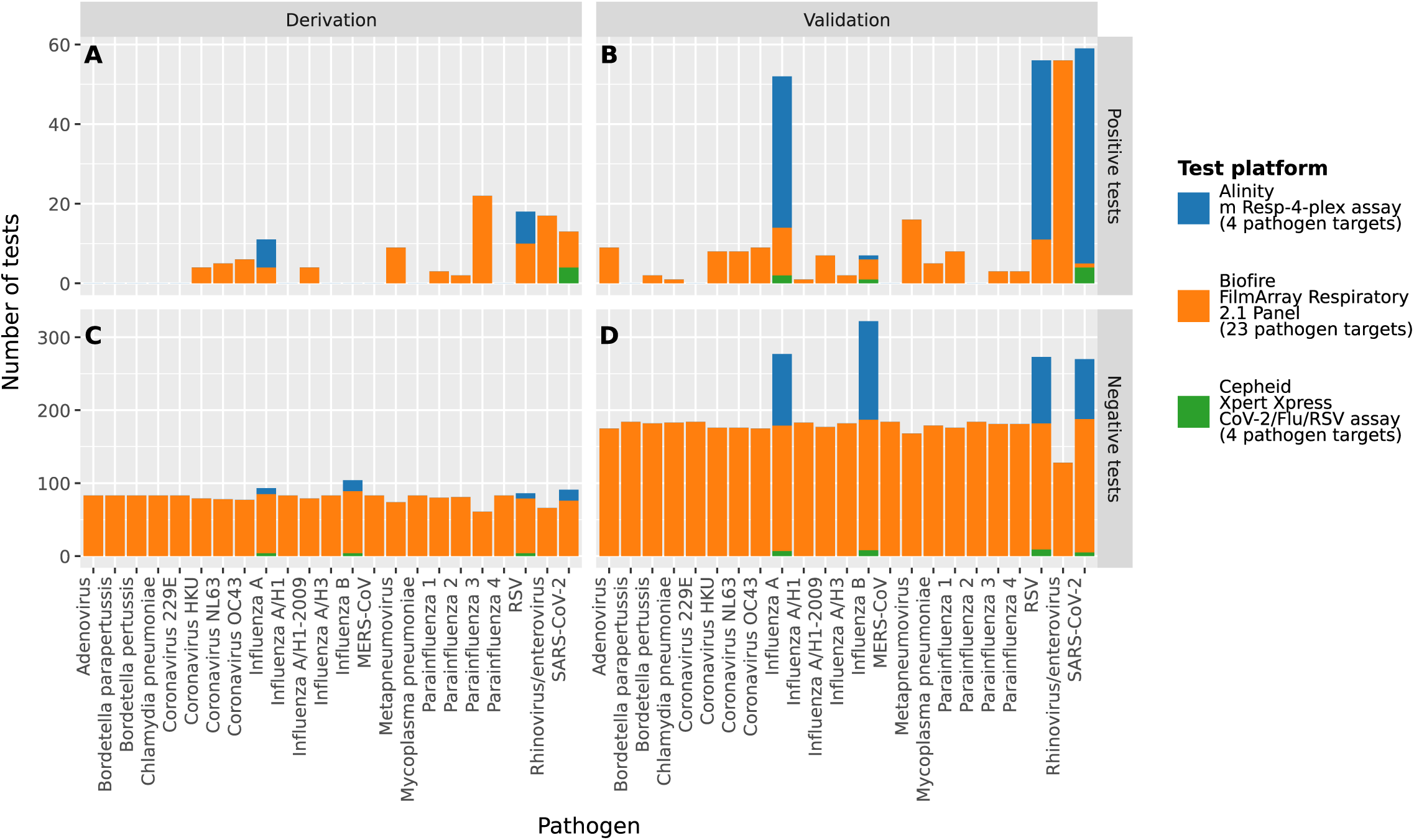
Pathogens identified (A, B) and not identified (C,D) on routine diagnostic testing in (A, C) derivation and (B, D) validation sets. Each sample could be positive for multiple pathogens, see **Table 1**.

### Derivation Set: Improved base-calling accuracy and read thresholds reduce false-positives from barcode crossover

In the derivation set, barcode crossover following standard high accuracy base-calling (HAC) led to sequencing reads being mis-attributed to another sample within a multiplexed pool (**Fig.S2**). The super accuracy base-calling (SUP) model demonstrated improved performance, reducing median reads assigned to unused barcodes from 0.1% (0.08%-0.11%) to 0.01% (0.01%-0.013%), and the number of sequences with ≥1 read mapping to a pathogen for which the sample was PCR-negative from 63/104 (62%) to 34/104 (33%). All 34 false-positives in SUP-called sequences occurred on sequencing runs which included another PCR-positive sample with a high number of reads for the pathogen detected, and were based on single read in 12/104 (12%) sequences. We therefore implemented a “≥2 reads” threshold for target pathogen detection (**Tab.2**). We found 56/104 (54%) derivation set sequences with ≥2 reads mapped to the pathogen identified by routine PCR testing and 18/104 (17%) with ≥2 reads mapping to a pathogen for which the sample was PCR-negative.

This “≥2 reads” quality threshold was also applied to sequencing batch positive and negative controls, and ICs (**Tab.2**). 2/6 (33%) derivation sequencing runs, containing 24/104 (23%) sequences, failed sequencing batch negative control thresholds (i.e. had ≥2 reads mapping to MS2 or one the three viral internal amplification controls). Among runs passing sequencing batch controls, 19/80 (24%) sequences failed ICs: 8/19 (42%) failed to detect two IC viruses and 11/19 (58%) failed to detect all three IC viruses (**Fig.3C**). In total, 61/104 (59%) derivation set sequences passed all negative and positive batch control and ICs and were used to establish bioinformatic thresholds for pathogen detection.

**Figure 3.**
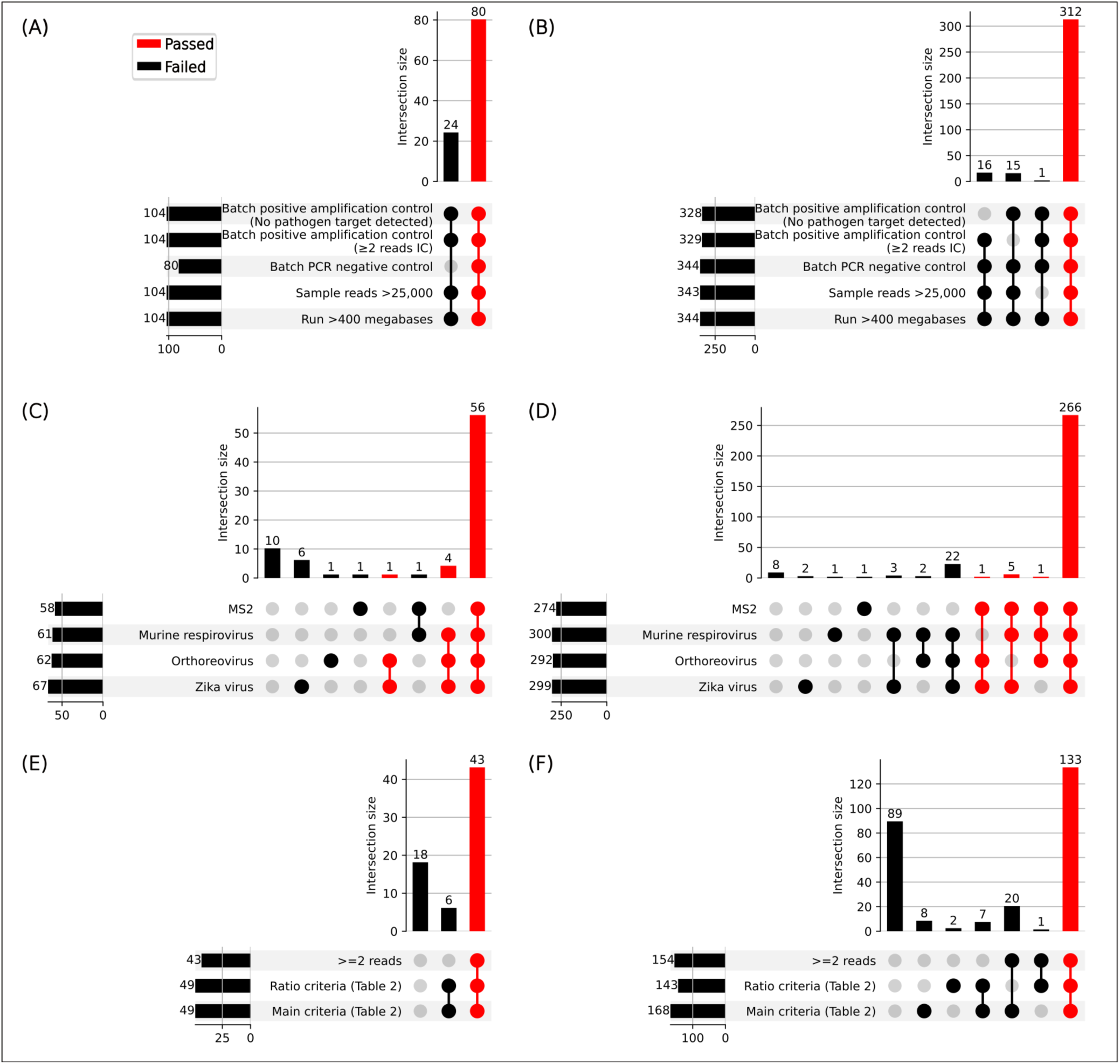
Batch control (A, B) and positive control (C, D) passed per sample and target detection (E, F) criteria passed for (A, C, E) derivation and (B, D, F) validation sets. Red indicates passed and black failed. RNA from murine respirovirus, orthoreovirus and Zika virus were spiked as cDNA synthesis and SISPA controls, MS2 phage was spiked as an extraction, cDNA synthesis and SISPA control. Within the derivation set MS2 was not required to pass QC due to poor performance of the laboratory MS2 stock; in both validation and derivation only detection of 2/3 virus spikes was required to pass QC.

### Derivation Set: Defining bioinformatic thresholds for pathogen detection

Within 61 derivation sequences passing QC, 67 pathogen targets were detected by routine PCR (i.e. gold-standard positives), and 1154 pathogen targets were not detected by PCR (i.e. gold-standard negatives) (**Tab.1**). We evaluated 28 bioinformatic metrics against true-positives and true-negatives to optimise detection (**Tab.S3**;**S4**), individually and in combination (**supplemental methods**). Many metrics and combinations had very similar performance (**Tab.S3;S4;S5**), and were highly collinear (**Fig.S3**). Maximising Youden’s index, and taking into account sequencing performance aspects (**Tab.S3**), we chose a final combination of three metrics encompassing coverage breadth, depth and the proportion of mapped reads, to address known sources of error in metagenomics.^23,24^ Specifically, a pathogen detection in sequence data required a) the percentage of the reference genome with coverage of ≥1 read was >0.25%, or b) the mean read depth (truncated at a maximum of 10 at all positions) across the entire genome (denoted “area under the genome”) was >0.003, or c) the number of reads in the sample mapping to the target reference genome as a percentage of pathogen reads on the run mapping to any pathogen reference genome was >0.006% (**Tab.2**).

Combining these three “OR” criteria with the “AND” criterion for ≥2 target pathogen reads led to 28/1929 false-positives including all sequences in the derivation set (including those not passing batch/internal QC control), and 4/1154 false-positives after restricting to sequences that passed QC checks. False-positives had proportionately lower “area under the genome” compared with the percentage of reads in the sample mapping to the target reference genome as a percentage of reads on the run mapping to any pathogen reference genome (**Fig.4A**). We therefore added a third “AND” “ratio criterion” for positivity, specifically that the ratio between these criteria should be >0.1 (**Tab.2, Fig.4A**).

**Figure 4.**
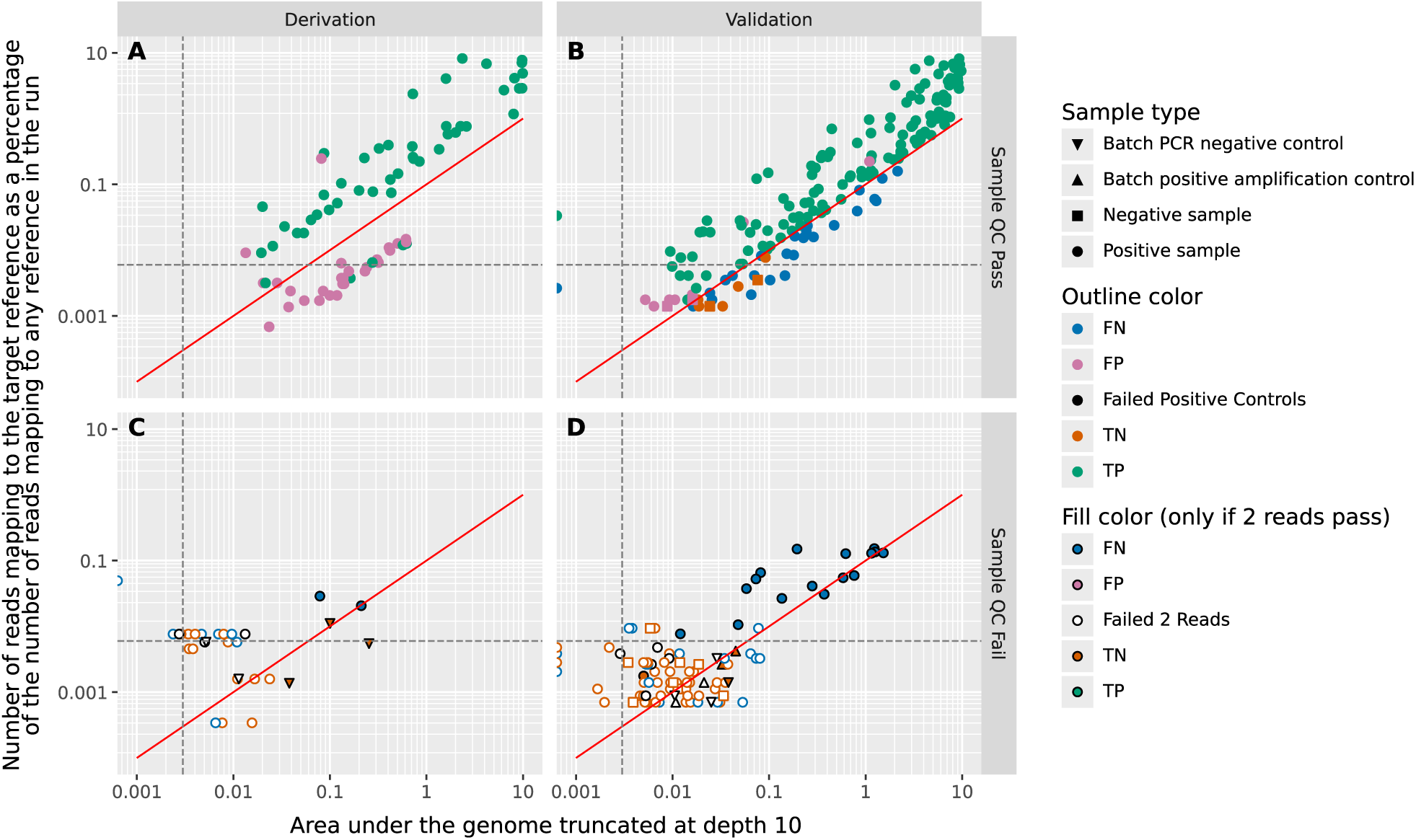
Positivity ratio criteria as developed in the derivation set (A,C) and tested in the validation set (B,D) Showing samples which passed sample QC thresholds (**Tab.2**) in panels A, B and samples which failed sample QC thresholds in panels C, D. The sample QC pass (panels A, B) includes samples that failed batch controls, to provide context for barcode crossover. In panel A classification of gold standard PCR-positives in the derivation set as true or false was based only on the number of reads and the main criteria (**Tab.2**), ie did not include the ratio criteria shown by the red line, which was developed from the derivation set. In panel B, the classification of gold standard positives in the validation set does include the ratio criteria as shown by the red line. Individual thresholds shown in grey dashed lines.

Hence the final pathogen detection criteria determined were: (percentage of reference genome with ≥1 read >0.25% OR “area under the genome” >0.003 OR percentage of reads mapping to reference genome>0.006%)) AND ≥2 target pathogen reads AND “ratio criterion”>0.1 (**Tab.2**).

43/67 (64%) PCR-positives in the derivation set were detected by these three criteria (**Fig.3E**): among 24 PCR-positives classified negative, 18/67 (27%) had no reads mapped to the target pathogen, and 6/67 (9%) had one read mapped to the target pathogen (failed the ≥2 reads threshold). 1151/1154 (99.7%) of the pathogens tested for, but negative by multiplex PCR, were not detected by sequencing. The three false-positive target detections, occurring in 2 of the 38 original samples, were SARS-CoV2 (two reads) and influenza A, further subtyped as H1-2009 (five reads; counted as two target detections).

### Validation Set: Diagnostic performance

To validate the performance of the derived thresholds, we used 344 independent upper respiratory tract samples with known multiplex PCR results (**Tab.1a**) from patients with a wide range of ages and inpatient and emergency locations (**Tab.1b)**. Samples were positive for diverse pathogens by PCR, dominated by RNA viruses with a small number of DNA viruses and bacteria (**Fig.2**).

Samples were extracted in 22 batches and sequenced on 11 runs (**Fig.5**). 2/22 (9%) batches (containing 31/344 (9%) samples) failed batch QC, with one batch amplification control sample failing all three IC viruses (<2 mapped reads), and one batch amplification control sample failing due to detection of reads from a target pathogen (**Fig.5**). 39 samples failed internal extraction or amplification QC, 38/39 (97%) failing to detect the MS2 extraction and amplification control; in addition, 2/39 (8%) did not detect two IC viruses and 9/39 (23%) failed to detect any IC viruses (**Fig.3D**). Overall, 273/344 (79%) respiratory samples in the validation set passed negative and positive control QC criteria.

**Figure 5.**
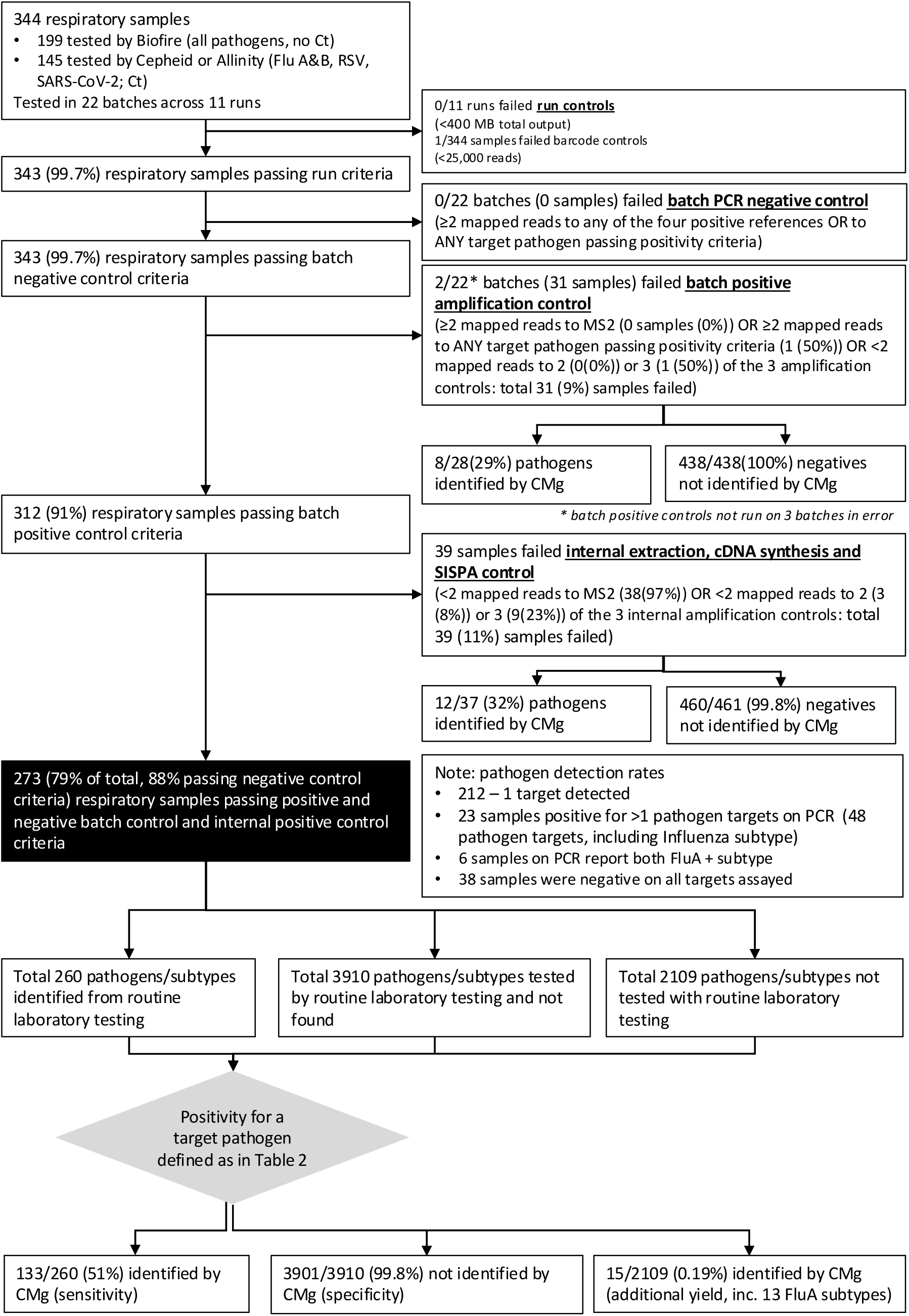
Validation set sample flow.

In these 273 samples, there were 260 pathogen gold-standard target detections by multiplex PCR, with 212 (78%) samples positive for a single pathogen, 21 (8%) positive for two pathogen targets (42 targets in total), 2 (1%) positive for 3 pathogen targets (6 targets in total) (**Tab.1**). Within these detections, six (2%) samples were positive for influenza A with additional detection of the H1 subtype (counted as 2 detections, following the FilmArray panel). 38/273 (14%) samples passing QC were PCR-negative for all pathogens.

133/260 (51%, 95%CI: 45-57%) PCR-positive results (i.e. gold-standard positives) were also detected by sequencing (**Tab.3**, **Figs3D, 4B**). Of samples with multiple pathogens targets found by PCR, 1/21 (4.8%) had two pathogens targets detected by sequencing, while 0/2 (0%) had three pathogen targets detected by sequencing. There were 127 false-negative results: 89 (34% of the 260 total results) had no reads mapped to the target pathogen, 17 (7% of the total) had only a single mapped read and therefore fell below the threshold designed to minimise barcode crossover, 20 (8%) had ≥2 reads and met at least one of the three main criteria but failed the ratio criteria (as defined in **Tab.2**), and 1 (0.4%) had ≥2 mapped reads and achieved the ratio criteria but did not meet any of the three main criteria (**Figs.3D, 4B**).

**Table 3.**
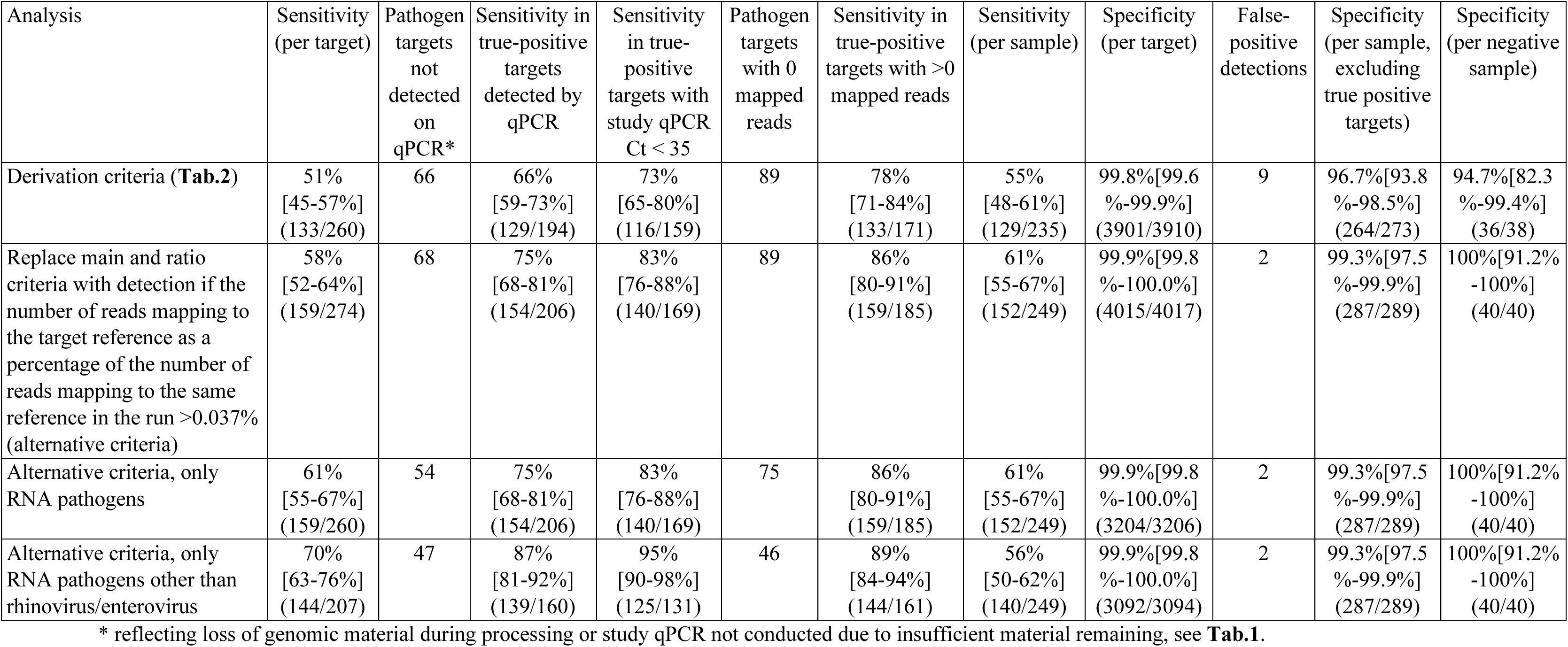
Sensitivity and specificity of different criteria for positivity in validation set.

3901/3910 (99.8%) targets negative by PCR (as tested in the original assays) were also not detected from sequencing (**Tab.3**); 38 samples were PCR-negative for all pathogens, of which 36 (95%) were also negative by sequencing. There were nine false-positive pathogen detections by sequencing, all potentially attributable to barcode crossover (**Fig.4B**): specifically a) four SARS-CoV2 detections with two reads each, all on a single run that contained a sample with early cycle SARS-CoV2 detection on PCR (Resp-4-Plex Cn=13.54; 51,158 reads); b) four coronavirus detections (3 HCoV-HKU1, 1 HCoV-OC43) with 2-38 reads from one run (Run 11) that contained a sample with 33,347 reads mapping to HCoV-HKU1 (PCR by FilmArray, no Cn/Ct reported); and c) one metapneumovirus detection with 79 reads from a run including two other metapneumovirus-positive samples with 48 and 810 reads (PCR by FilmArray).

The ratio criterion performed poorly in the validation set compared with the derivation set, resulting in 20 false-negatives solely from applying this criterion (**Figs.3D, 4B**). We therefore compared the main and ratio criteria (**Tab.2**) with an alternative criterion which had relatively similar performance to these criteria in the derivation set (**Tabs.S3, S5**). This criterion defined detection by the number of reads in the sample mapping to the target reference genome as a percentage of reads on the run mapping to the specific target genome (rather than all reference genomes) being >0.037%. Replacing the main and ratio criteria with this alternative criterion (but retaining the ≥2 reads criterion) increased the number of samples passing batch controls from 260 to 274, increased sensitivity to 58% (95% CI 52-64%; 159/274), and improved specificity to 99.9% (95% CI 99.8-100%; 4015/4017) with only 2 false-positive detections (**Tab.3, Fig.S4**), the metapneumovirus and coronavirus OC43 detections.

### Performance by pathogen

While overall sensitivity using pre-specified criteria was 51%, assay performance varied substantially by pathogen. Sequencing detected fourteen different RNA viruses in samples passing QC, but no bacteria or DNA viruses. *Chlamydia pneumoniae* was detected (3 reads) in one sample whose positive controls failed (**Fig.6A**). Among pathogen targets with more than 20 PCR-positive results, sensitivities using pre-specified criteria were: RSV 85% (95% CI: 75-95%); SARS-CoV-2 65% (95% CI: 53-78%); influenza A 58% (95% CI: 40-75%), and rhinovirus/enterovirus 19% (95% CI: 8-29%), with small improvements using the alternative criteria for all except SARS-CoV-2 (**Fig.6B**).

**Figure 6.**
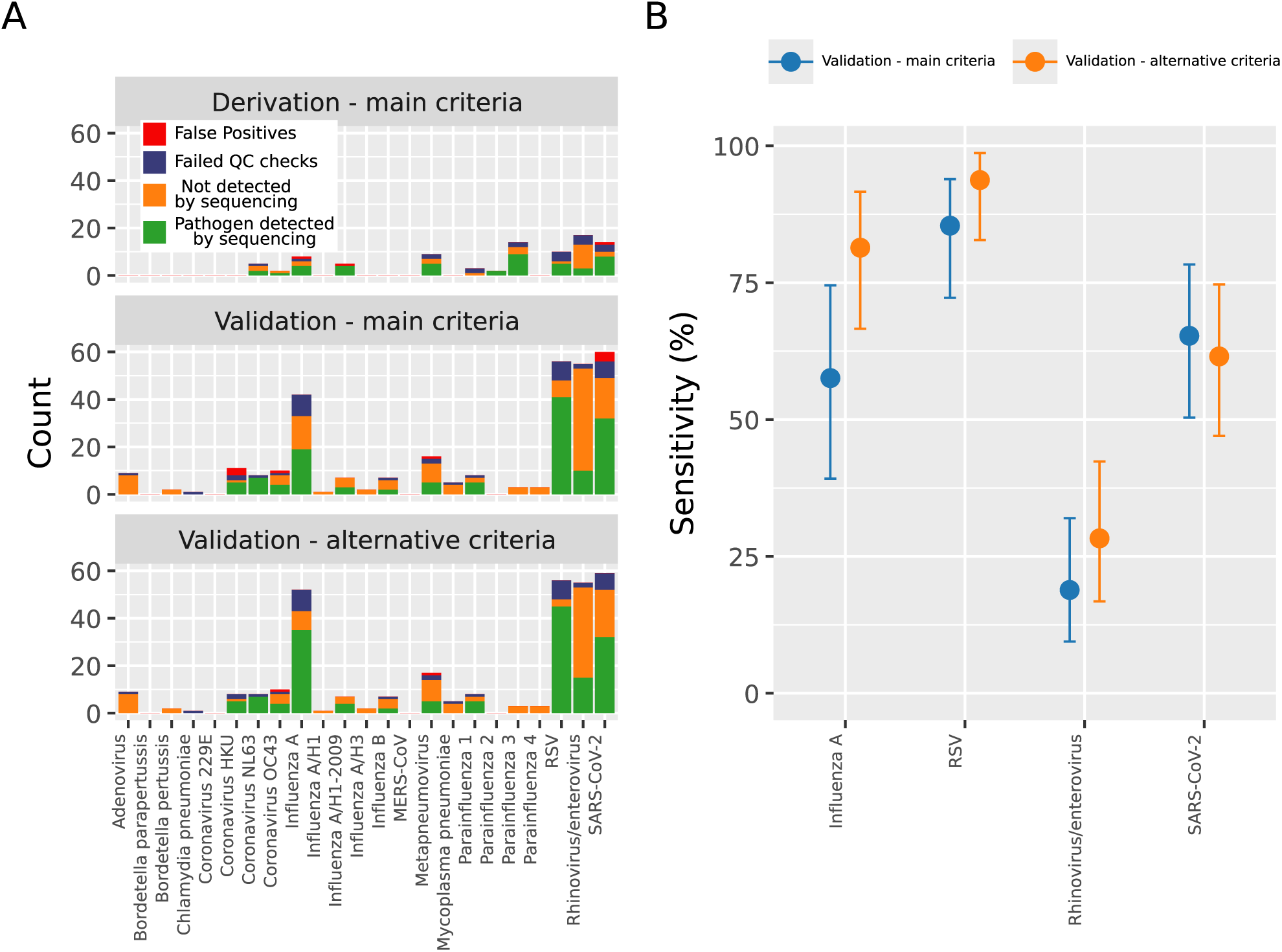
Pathogen-specific detections (A) and sensitivity (if >20 gold-standard detections) (B) in the derivation and validation sets. 95% confidence intervals calculated using Clopper-Pearson interval based on Beta distribution.

The sample workflow was designed to optimise RNA pathogen detection. Excluding DNA pathogen targets (adenovirus and bacteria) from the evaluation modestly increased sensitivity using the alternative criteria to 61% (159/260) (95% CI 55-67%) (**Tab.3**). Enterovirus and rhinovirus infections were frequently at low viral loads (**Fig.S5B**), of unclear significance;^25^ removing these two targets increased overall sensitivity using the alternative criteria further to 70% (145/207) (95% CI 63%-76%) (**Tab.3**).

### CMg provides greater taxonomic resolution and additional target detection

Ten rhinovirus/enterovirus FilmArray positive samples were further identified by sequencing as Rhinovirus B3 (n=6), Rhinovirus C (n=1), Rhinovirus A89 (n=1), and Enterovirus A (n=2). All 41 samples with RSV detected by sequencing were further identified as either RSV-A (n=22) or RSV-B (n=19). In addition, 13 samples with influenza A detected on 4-plex PCR were subtyped with sequencing: 12 as influenza A/H1-2009 and one as influenza A/H3.

We detected two pathogens (rhinovirus and coronavirus OC43) by CMg sequencing that were not assayed in the original PCR test in 2/111 (2%) samples originally tested by 4-plex. These were not on runs with other strong positives, representing plausible additional detection.

### Performance by viral load (Cn,Ct)

We compared PCR Cn/Ct values from the original Resp-4-Plex/Xpert platforms with Ct values from study-specific qPCR following ss cDNA synthesis (**Fig.S5**). Study qPCR Ct values were consistently 5-10 cycles higher than original Cn/Ct values, potentially due to loss of material during processing and/or to the use of different assays.

Samples with higher Ct values from either assay generated fewer sequencing reads as expected (**Fig.7**). When using the alternative detection criteria and restricting analysis to samples with detectable pathogen by qPCR, sensitivity increased to 75% (95% CI, 68-81%), and increased further to 83% (95% CI, 76-88%) when restricting to qPCR Ct <35 (**Tab.3**).

**Figure 7.**
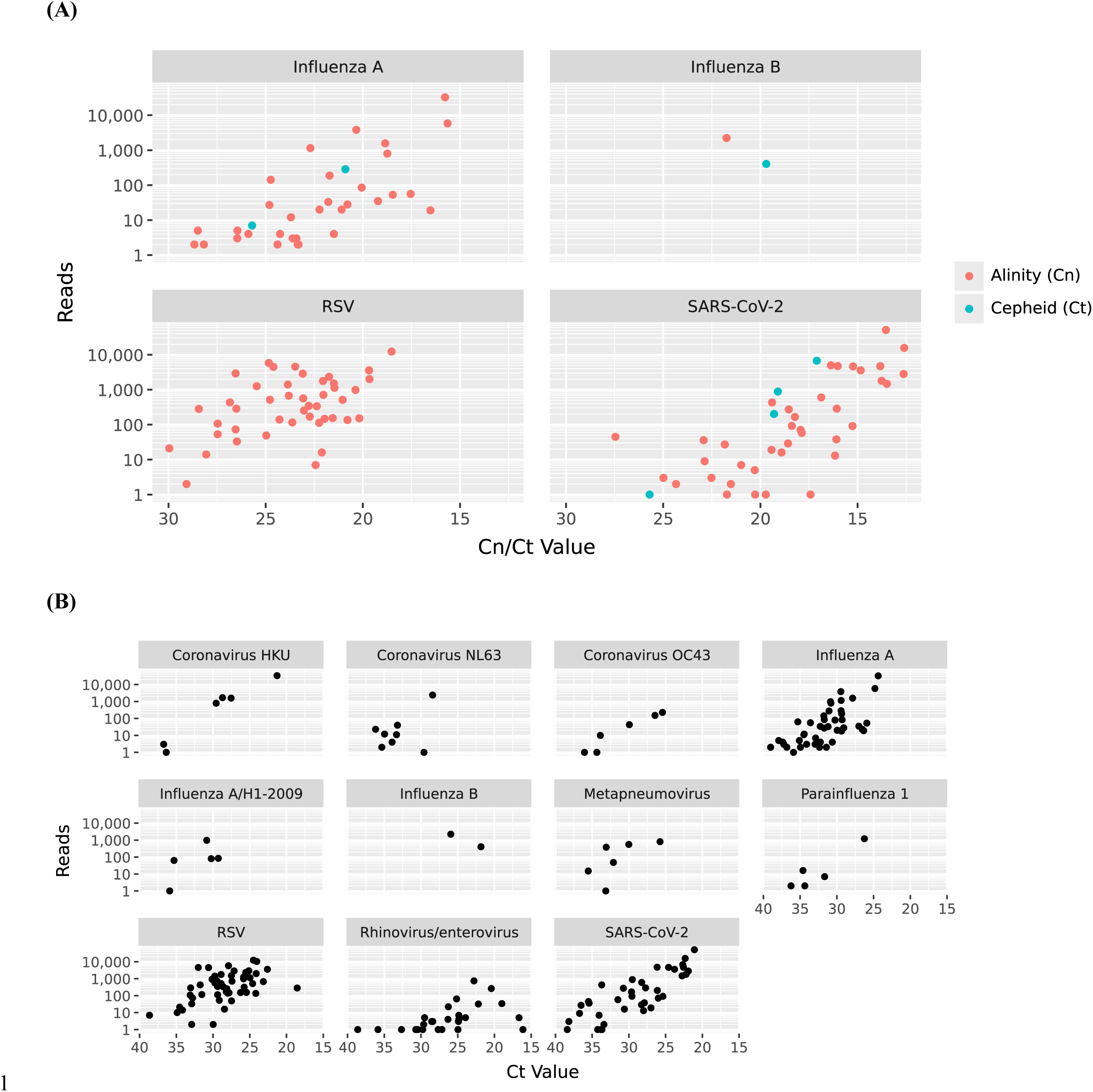
Association between number of mapped reads and Cn/Ct values from original assay (A) or study qPCR (B)

### Sequencing assay costs

The sequencing and consumables costs were calculated based on multiplexing 32 clinical samples on each sequencing run at £102.41/sample (**Tab.S6**). Sequencing platform costs added a further £9.61/sample. The analysis pipeline can be performed using GridION onboard computing capacity; no additional server or compute costs were therefore incurred. In addition to £112.02 sequencing and infrastructure costs, each sample required 70 minutes of labour.

## Discussion

Here, we developed and assessed the performance of an end-to-end metagenomic workflow to detect a range of RNA respiratory viruses. By applying stringent quality thresholds, we achieved high specificity both for target pathogens (99.8%) and at an individual sample level (>96%). Sensitivity using pre-defined thresholds in a large validation set was modest compared to multiplex PCR, being 51% overall, ranging from 85% for RSV to 19% for rhino/enterovirus. This could be improved to 83% by limiting analysis to RNA viruses with Ct <35 and using post-hoc exploratory bioinformatic criteria. The final cost was £112/sample, achieved by 32-multiplexing across one flow cell. While cost compares favourably with Filmarray, and the sequencing cost was 45% less than an Illumina-based respiratory virus sequencing assay,^17^ the hands-on and labour time of the assay was substantial (estimated 70 minutes/sample).

The ultimate goal of the assay was agnostic pathogen detection: however, a necessary first step was reliable detection of known targets from routine testing. We therefore developed a mapping-based analysis pipeline, the most sensitive approach to demonstrate performance against known targets.^22^ We validated this workflow against PCR targets using prospectively collected samples including 19/23 multiplex-PCR targets, substantially broader than the 4-8 targets validated in other RNA virus CMg workflows to date.^16,17^

The varying sensitivities obtained highlight challenges of CMg for detecting infections with high host and low pathogen biomass (as from nasopharyngeal swabs). Despite human cell depletion and RNA amplification, only 66% of samples yielded sequence reads for the detected PCR target, and 20% of gold-standard detections were not found by study qPCR. Such relatively low yield of pathogen reads has also been observed in other respiratory CMg workflows. One workflow required only 3 non-overlapping pathogen reads from a minimum of 5,000,000 per sample,^17^ while two studies reported a single positive read on ONT sequencing as the threshold for virus detection.^11,16^ Our results demonstrate that the consequence of relaxed thresholds is false-positive detections, a particular danger with imperfect barcode de-multiplexing.

The assay performed particularly poorly for rhinovirus/enterovirus. Genome size (7kb vs 13-30kb other viruses) may contribute to this poor sensitivity (**Tab.S2**). Additionally, differences between study sequences and the references used for mapping (**Tab.S2**) were greatest for rhinovirus/enterovirus (**Fig.S6**), suggesting that species diversity might also affect performance.

Our findings contrast with a recent UK respiratory CMg study, reporting sensitivity for viral detection of 89% (95%CI: 71-98),^14^ likely for multiple reasons. Alcolea-Medina et al sampled patients in critical care, with >85% of viral detections from lower respiratory tract samples, where common respiratory viruses have higher viral loads.^26,27^ Moreover, sparse host material in BAL samples allowed investigators to omit host depletion steps.^14^ Finally, the study multiplexed ≤5 samples, and 66% of included samples had no respiratory viruses detected by PCR testing. The lower risk of barcode cross-over allowed only a single read to indicate pathogen detection,^11^ albeit at greater sequencing costs.

RNA extraction followed by SISPA, as employed here, has high hands-on time and requires more specialised laboratory practitioners than commercial multiplex-PCRs. These technical aspects and the modest sensitivity of the workflow mean that it will not be scalable for routine diagnosis. However, metagenomic workflows using established laboratory methods and limited specialised equipment may be valuable for pandemic preparedness, since RNA viruses are likely candidates for emerging pathogens.^28–30^

CMg undoubtedly has diagnostic potential. However, with a laboratory workflow optimised for RNA extraction, and bioinformatics optimised to stringent recognition of sample and barcode cross-over, this study demonstrates the technical limitations of high-throughput metagenomic sequencing for routine viral diagnostics in upper respiratory tract samples with high host and low pathogen biomass. Its improved performance at higher viral loads demonstrates the importance of identifying the most valuable use cases for CMg.

## Data Availability

Code is available at https://github.com/oxfordmmm/AD_analysis Raw sequencing data available in the ENA at https://www.ebi.ac.uk/ena/browser/view/PRJEB102181

## Data Sharing

Deidentified patient data is included in supplementary material. All sequence data are available at the ENA (project number PRJEB102181).

## Code Availability

https://github.com/oxfordmmm/AD_analysis/tree/v1.0

## Contributions

Conception or design; N.D.S, K.D, D.E, A.S.W, B.C.Y. Acquisition, analysis or interpretation of data; N.D.S, K.D, K.M.V.H, M.C, M.P, V.D, T.P.Q, N.S., D.E, P.B., A.S.W, B.C.Y. Accessed and verified the data: N.D.S and B.C.Y. Drafted the work or substantively revised it: N.D.S, P.B., A.S.W, B.C.Y. All authors have had full access to the study data and accept responsibility to submit for publication

## Acknowledgements

This project was funded by the NIHR Biomedical Research Centre, Oxford, and supported by the NIHR Health Protection Research Unit in Healthcare Associated Infections and Antimicrobial Resistance at Oxford University in partnership with the UK Health Security Agency (UKHSA) (NIHR207397). The views expressed are those of the authors and not necessarily those of the NHS, the NIHR, the Department of Health and Social Care or UKHSA. ASW is an NIHR Senior Investigator. For the purpose of Open Access, the author has applied a CC BY public copyright licence to any Author Accepted Manuscript (AAM) version arising.

## Ethics declarations

The use of surplus material from respiratory samples processed by the Oxford University Hospitals NHS Foundation Trust Clinical Microbiology laboratory, and use of de-identified patient information from these samples, without patient-level consent, was approved by the Queens Square NHS Research Ethics Committee (REC), London, reference 17/LO/1420, as part of work on the development of rapid diagnostics.

