## Supplementary materials for "Clinical validation of a novel metagenomic nanopore sequencing method for detecting viral respiratory pathogens: diagnostic accuracy study"

| Contents | Page |
|---|---|
| **STARD checklist** | 2 |
| **Supplementary Methods** | 3-7 |
| **Supplementary Figures**<br>Table S1 Targets in multiplex PCR performed in routine testing<br>Table S2 Reference genomes used in the mapping pipeline<br>Table S3 Sensitivity and specificity of 8 different individual metrics to identify target pathogens in the derivation and validation sets<br>Table S4 Sensitivity and specificity of 20 other individual metrics considered for target pathogen identification in the derivation set<br>Table S5 Sensitivity and specificity of combinations of 2-8 of the 8 prioritised metrics to identify target pathogens in the derivation set<br>Table S6 Assay costs<br>Table S7 Sample testing and sequencing data (separate file) | <br>8<br>9<br>11<br>12<br>13<br>14 |
| **Supplementary Tables**<br>Figure S1 Total sequencing data yield and total number of reads per sample for derivation and validation sets<br>Figure S2 Barcode cross-over in derivation set (before and after super accuracy basecalling) and in validation set (with super accuracy basecalling)<br>Figure S3 Distribution of 8 key metrics in the derivation set<br>Figure S4 Positivity detection using alternative criteria in the derivation set (A) and validation set (B)<br>Figure S5 Association between Cn/Ct values from routine diagnostic PCR assay (Resp-4-Plex/Xpert) and study qPCR (A), distribution of Cn/Ct values from both (B), and percentage of samples with target detected by study qPCR (C).<br>Figure S6 Mapping quality scores from aligned reads by the percentage difference to the reference genomes | <br>15<br>16<br>17<br>18<br>19<br>21 |

#### STARD Checklist

| Section & Topic | No | Item | Reported on page # |
|---|---|---|---|
| **TITLE OR ABSTRACT** | | | |
| | **1** | Identification as a study of diagnostic accuracy using at least one measure of accuracy (such as sensitivity, specificity, predictive values, or AUC) | p2 |
| **ABSTRACT** | | | |
| | **2** | Structured summary of study design, methods, results, and conclusions (for specific guidance, see STARD for Abstracts) | p2 |
| **INTRODUCTION** | | | |
| | **3** | Scientific and clinical background, including the intended use and clinical role of the index test | p4 |
| | **4** | Study objectives and hypotheses | p4 |
| **METHODS** | | | |
| *Study design* | **5** | Whether data collection was planned before the index test and reference standard were performed (prospective study) or after (retrospective study) | p5 |
| *Participants* | **6** | Eligibility criteria | p5 |
| | **7** | On what basis potentially eligible participants were identified (such as symptoms, results from previous tests, inclusion in registry) | p5 |
| | **8** | Where and when potentially eligible participants were identified (setting, location and dates) | p5,9 |
| | **9** | Whether participants formed a consecutive, random or convenience series | p5 |
| *Test methods* | **10a** | Index test, in sufficient detail to allow replication | p5-6<br>Supp p3-6 |
| | **10b** | Reference standard, in sufficient detail to allow replication | p4 |
| | **11** | Rationale for choosing the reference standard (if alternatives exist) | p5 |
| | **12a** | Definition of and rationale for test positivity cut-offs or result categories of the index test, distinguishing pre-specified from exploratory | p6-9 |
| | **12b** | Definition of and rationale for test positivity cut-offs or result categories of the reference standard, distinguishing pre-specified from exploratory | p5 |
| | **13a** | Whether clinical information and reference standard results were available to the performers/readers of the index test | p5 |
| | **13b** | Whether clinical information and index test results were available to the assessors of the reference standard | p5 |
| *Analysis* | **14** | Methods for estimating or comparing measures of diagnostic accuracy | p4 |
| | **15** | How indeterminate index test or reference standard results were handled | Nil indeterminate |
| | **16** | How missing data on the index test and reference standard were handled | Nil missing |
| | **17** | Any analyses of variability in diagnostic accuracy, distinguishing pre-specified from exploratory | p10-11 |
| | **18** | Intended sample size and how it was determined | p5 |
| **RESULTS** | | | |
| *Participants* | **19** | Flow of participants, using a diagram | Fig.5 |
| | **20** | Baseline demographic and clinical characteristics of participants | Tab.1 |
| | **21a** | Distribution of severity of disease in those with the target condition | Not relevant |
| | **21b** | Distribution of alternative diagnoses in those without the target condition | Not relevant |
| | **22** | Time interval and any clinical interventions between index test and reference standard | p5 |
| *Test results* | **23** | Cross tabulation of the index test results (or their distribution) by the results of the reference standard | Tab.3 |
| | **24** | Estimates of diagnostic accuracy and their precision (such as 95% confidence intervals) | Tab.3 |
| | **25** | Any adverse events from performing the index test or the reference standard | Nil |
| **DISCUSSION** | | | |
| | **26** | Study limitations, including sources of potential bias, statistical uncertainty, and generalisability | p13-14 |
| | **27** | Implications for practice, including the intended use and clinical role of the index test | p14 |
| **OTHER INFORMATION** | | | |
| | **28** | Registration number and name of registry | Not done |
| | **29** | Where the full study protocol can be accessed | n/a |
| | **30** | Sources of funding and other support; role of funders | p2, p15 |

### Supplementary Methods

#### Sample collection and processing

Nasopharyngeal samples collected with flocked swabs were placed in viral transport medium (VTM) (UTM (Copan, Italy) or Sigma-Virocult (MWE, UK)) for routine clinical testing in the Microbiology Laboratory, Oxford University Hospitals NHS Foundation Trust (Oxford, UK). Samples surplus to clinical testing were retrieved. Inclusion criteria were: (i) nose and throat swabs, (ii) <7 days since patient sampling, iii) pathogen detected at cycle threshold (Ct) or cycle number (Cn) value ≤30 for Resp-4-Plex (Abbott, USA) and Xpert assays (Cepheid, USA) or sampled tested by BIOFIRE FilmArray Respiratory 2.1 Panel (Biomerieux, France); the latter assay does not report a Ct value.

For the derivation set, samples were selected to cover the broadest range of pathogen detections by PCR between March and July 2024. For the validation set, all positive samples meeting inclusion criteria for the winter 2024/25 respiratory season, i.e. between October 2024 and March 2025, were retrieved, until a threshold of n=60 positives per virus was reached (met for RSV, SARS-CoV-2 and human rhino/enterovirus). Approximately one FilmArray negative sample was collected for every eight positives (by any method).

Clinical laboratory results were confirmed by retrieving data from the Laboratory Information Management System. This included date collected, test type, pathogen result, and Cn/Ct value if available.

**Host Nucleic Acid Depletion and Virus Enrichment**
Prior to RNA extraction, samples were pre-treated to reduce host nucleic acid and enrich for virus particles. Swab-containing VTM was vortexed (30 seconds) and 1mL removed. Samples were clarified by low speed centrifugation (3,200g, 30 min), pelleting intact human cells and debris. The supernatant was transferred to a fresh lo-bind Eppendorf tube and free nucleic acid was digested using M-SAN HQ (1000U) (ArcticZymes Technologies, Tromsø, Norway) with final $MgCl_2$ and Tris HCl pH 8.0 concentrations adjusted to 10mM and incubation at 37°C for one hour. 500µl Intact Virus Precipitation Reagent (Thermo Fisher Scientific, Waltham, MA, USA) was added, mixed by pipetting and incubated overnight (4°C). Precipitants were collected by high speed centrifugation (13,200g, 30 min) and the supernatant was discarded. Phosphate buffered saline (PBS, Sigma Aldrich, USA) (20µl) and MS2 extraction control (10µl) (Thermo Fisher Scientific) were added and the pellet resuspended gently by pipetting, prior to RNA extraction.

**RNA Extraction**
Samples (30µl) were extracted using the Direct-zol RNA Microprep kit (Zymo Research) according to the manufacturer instructions, including the optional on-column DNase treatment. RNA was eluted in DEPC treated water (10µl) (Ambion, Thermo Fisher Scientific).

**cDNA Synthesis and SISPA**
First strand cDNA was synthesized using the Maxima H Minus 1$^{st}$ Strand cDNA synthesis kit with ds DNase (Thermo Fisher Scientific), in a 20µl reaction volume, according to the manufacturer's protocol, with the following modifications. Extracted RNA (8µl) was treated with DNAse, as recommended, then spiked with three reverse transcription/amplification controls (1µl each, containing approximately $10^4$ genome copies each of Zika virus (ATCC-VR-1838DQ), murine respirovirus (ATCC-VR-907DQ) and orthoreovirus (ATCC-VR-824DQ) (ATCC Manassas, Virginia, USA). cDNA synthesis was primed randomly using the 3' terminal $N_9$ segment of custom oligonucleotide primers (5'-GAT-GAT-AGT-AGG-GCT-

TCG-TCA-CNN-NNN-NNN N-3'; Integrated DNA Technologies, Leuven, Belgium), final concentration 5µM. This incorporated the SISPA primer sequence (5'-GAT-GAT-AGT-AGG-GCT-TCG TCA-3') into the first strand cDNA. Following the recommended denaturation at 65°C for 5 min, 5x RT buffer and Maxima H minus Enzyme Mix were added and primer annealing, reverse transcription and inactivation took place under recommended conditions, 25°C for 10 min, 50°C for 30 min and 85°C for 5 min. Following reverse transcription, cDNA fragments >~100 nucleotides were purified using SPRI beads (AMPure XP beads, Beckman Coulter, Indianapolis, Indiana, USA) with two 70% ethanol washes and elution in 16µl DEPC-treated water (Ambion, Thermo Fisher Scientific). A 3µl cDNA aliquot was reserved for separate study qPCR, and 11µl taken forward into second strand cDNA synthesis.

Second strand cDNA was synthesized using Sequenase™ V2.0 DNA Polymerase (Thermo Fisher Scientific) and the same custom oligonucleotide primer, ((5'-GAT-GAT-AGT-AGG-GCT-TCG-TCA-CNN-NNN-NNN N-3') randomly priming second strand synthesis and incorporating the SISPA oligonucleotide primer. Reaction conditions were as described previously[1] comprising 11µl ss cDNA, 4µl 5x sequenase buffer, and 1µl oligonucleotide primer (40 µM). Samples were denatured at 97˚C for 2 minutes and cooled to 4˚C, then 1µl DTT (100mM), 1µl dNTPs (10mM) and 1.8 µl DEPC-treated water were added, followed by incubation at 37˚C for 8 minutes. A second identical incubation was performed following the addition of 0.55µl sequenase dilution buffer and 0.2 µl Sequenase. Following SPRI bead purification, dsDNA >100 bp was eluted in 22µl nuclease free water, 20µl of which was amplified by SISPA PCR.

Each reaction (50µl) comprised 20µl ds cDNA, 5µl SISPA 1 oligonucleotide (5'-GAT-GAT-AGT-AGG-GCT-TCG-TCA-3') (20 µM) and 25µl NEBNext® Ultra™ II Q5® Master Mix (New England Biolabs, Ipswich, Massachusetts, USA). A no template negative control replacing cDNA with 20µl DEPC-treated water was included at this stage. Amplification conditions were denaturation at 98°C for 30 seconds (1 cycle), followed by 30 cycles of denaturation at 98°C for 10 seconds, annealing 64°C for 30 seconds, and extension 72°C for 1 min 30sec (30 cycles), and a final extension of 72°C for 2 minutes. A final SPRI beads purification was followed by elution in 15µl DEPC-treated water. The amplification products were quantified by Qubit fluorometer using the dsDNA 1X BR Assay (Thermo Fisher Scientific).

**Library preparation and sequencing**
Libraries were prepared according to the Rapid Sequencing V14-Amplicon Sequencing protocol from ONT. Samples were sequenced using Rapid Barcoding Kit V14 (SQK-RBK114.96; ONT, Oxford, UK) on R10.4.1 Flow Cells (ONT). A maximum of 36 samples were run on a single flow cell (32 clinical samples plus 4 positive/negative controls), and were run for 72 hours (with SUP base calling enabled for the validation set).

#### qPCR of extracted cDNA

During cDNA synthesis, 3µl of single-stranded cDNA was reserved for quantification. Samples were tested by qPCR assay targeting the respiratory virus(es) detected on gold-standard testing. Gold-standard RSV positives underwent separate qPCR for RSV-A and RSV-B. Gold standard rhinovirus/enterovirus positives underwent separate qPCR for enterovirus and rhinovirus. Due to limited cDNA availability, only 3 qPCR reactions were possible per sample. Samples with gold standard detection of >1 pathogen target were prioritised for quantification with enterovirus and rhinovirus if rhinovirus/enterovirus was detected, followed by other RNA viruses. Reaction conditions were as recommended by the manufacturer; 0.5µl TaqMan Microbe Detection Assay (Thermo Fisher Scientific), 5µl TaqMan Fast Advanced Master Mix (Thermo Fisher Scientific), 3.5µl $H_2O$ and 1µl cDNA. Thermocycling was performed on the Quant Studio 5 PCR machine with the following conditions; initial denaturation at 95°C for 20 seconds followed by 40 cycles of denaturation at 95°C for 1 second and annealing/extension at 60°C for 20 seconds. Standard curves for quantification were generated by serially diluting True Mark Respiratory Panel 2.0 Amplification Control (Thermo Fisher Scientific) for a dilution range between $10^5$ to $10^1$ copies per reaction. Standard curves were included on every qPCR reaction plate in triplicate. Quantification and Ct values were determined using the Design and Analysis Software (Thermo Fisher Scientific) with the "Quantification-TaqMan, Standard Curve" settings enabled.

#### Bioinformatic analysis

Raw ONT sequencing reads were basecalled on a GridION Mk1 with using Dorado (ONT) with the current basecalling models at the time of sequencing: dna_r10.4.1_e8.2_400bps_hac@v4.3.0 (derivation set) or dna_r10.4.1_e8.2_400bps_sup@v4.3.0 (validation set). The derivation set ONT sequences were subsequently basecalled again with the then current standalone model dna_r10.4.1_e8.2_400bps_sup@v5.0.0 model and hardware (Nvidia RTX2080 GPU and Intel(R) Xeon(R) Gold 5217 CPU @ 3.00GHz). It was not possible to re-call the validation set with this model as the ONT output data was changed in July 2025 and the relevant pod5 files were not retained by default.

Human host reads were filtered from the output fastq files before downstream analysis using kraken2 and the k2_pluspf_20240605 database (https://benlangmead.github.io/aws-indexes/k2). Further read removal was performed using Deacon[2] with default settings before uploading to the ENA (project number PRJEB102181).

The analysis workflow is provided on https://github.com/oxfordmmm/AD_analysis (v1.0). Read and base numbers for each run and sample were calculated with SeqKit[3] v2.9.0. SISPA primers were identified with BLASTn[4] v2.16.0+ (using parameters: -task blastn-short, word_size 7, gapopen 2, max_target_seqs 10000000) and trimmed using the trim_primer.py script. Reads with more than 10 SISPA primers were removed before mapping. Reads were mapped using minimap2[5] v2.22-r1101 (using parameters: -ax map-ont -N 1000) to a curated composite reference fasta file including representative genomes from a range of respiratory viral and bacterial pathogens (**Tab.S5**). Due to non-specific mapping to the larger genomes (**Fig.S6**), reads mapping to the bacterial genomes with a mapQ score <40 were excluded. This was not applied to the reads mapping to the viral genomes.

We considered 28 potential bioinformatic metrics created from mapping the raw sequencing reads to the composite references in samples passing QC in the derivation set (**Tab.S2;S3**). Sequencing coverage metrics used for species classification were calculated using samtools[6] v1.21 and the coverageStats.py script. Metrics considered covered the following aspects

- Coverage: at 1x, 3x, 5x or 10x depth, in absolute number of bases and as a percentage of reference length (ie normalised)
- Read-depth coverage across the whole genome ("area under the genome") and divided by the reference length: at overall, and truncating read depth to 5x or 10x to reduce the impact of mis-mapped spikes of coverage

- Mean read-depth coverage across the genome with read depth at least one, excluding from the denominator regions with no coverage, overall and truncating to 5 or 10 read depth.
- Bases mapped to the reference, absolute and as a percentage of reference length (ie normalised)
- Reads mapped to reference, excluding multiple supplementary alignments on the same reference but including secondary alignments on different references, overall, and restricting to reads with alignment length ≥200, ≥300, ≥400, or ≥500.
- Reads mapped to a target in a sample as a percentage of the total number of reads in the run, the total number of reads in the run mapping to any reference or the total number of reads in the run mapping to references from the target species (the last will be 100% where there only one reference from a species and only one positive for that species on the run and no false-positives on the run).
- Reads mapped to a target in a sample as a percentage of the total number of reads in the sample mapping to references from the target species (100% where there only one reference from a species and only one positive for that species on the sample).

We excluded the most collinear based on Spearman correlations, leaving eight metrics for detailed consideration (**Tab.S2**, **Fig.S3**).

#### Statistical analysis

Area under the receiver operating curves (AUROC) for the eight metrics undergoing detailed consideration ranged from 0.854-0.864 (**Tab.S2**). Detection thresholds for each metric were determined using Youden's index performed on all QC passing samples in the derivation set. The gold standard was the multiplex PCR results (FilmArray, Resp-4-Plex, Xpert, **Tab.1**). Each sample contributed N observations, one for each of the potential pathogen targets that could have been detected by the PCR test used (FilmArray 23, Resp-4-Plex/Xpert 4). For influenza A, FilmArray reports positive/negative overall, and then can report none or one of three subtypes if positive. These subtypes were only included as targets in analysis if detected by FilmArray. For sequencing detection, the subtype (H1, H1-2009, H3) was determined by the highest number of mapped sequencing reads, and Influenza A was counted as positive if any subtype was positive.

We then calculated the sensitivity and specificity of all 255 combinations of detection based on sequences meeting any one to eight of these metrics (**Tab.S4**). The maximum sensitivity of 0.731 was achieved by more than 30 combinations of three metrics (**Tab.S4**). We therefore selected a combination which covered three different aspects of sequencing performance, namely coverage breadth, depth of the mapped reads and the mapped reads as a percentage of reads mapping to any reference in the run.

AUROC analyses for each metric were performed using Python 3.10.4 and Sklearn[7] v1.1.1. 95% confidence intervals were calculated using binomial proportion with Clopper-Pearson exact interval using statsmodels[8] v0.15.0.

**Table S1. Targets in multiplex PCR performed in routine testing**

| | **Alinity Resp-4-plex assay** | **Xpert Xpress CoV-2/Flu/RSV plus** | **BIOFIRE FilmArray Respiratory 2.1 panel** |
|---|---|---|---|
| **RNA Virus** | Severe acute respiratory syndrome coronavirus 2 (SARS-CoV-2)<br>Influenza A virus<br>Influenza B virus<br>Respiratory syncytial virus | Severe acute respiratory syndrome coronavirus 2 (SARS-CoV-2)<br>Influenza A virus<br>Influenza B virus<br>Respiratory syncytial virus | Severe acute respiratory syndrome coronavirus 2 (SARS-CoV-2)<br>Coronavirus 229E<br>Coronavirus HKU1<br>Coronavirus NL63<br>Coronavirus OC43<br>Middle East respiratory syndrome-related coronavirus (MERS-CoV)<br>Human rhinovirus/enterovirus<br>Influenza A virus<br>Influenza A virus A/H1<br>Influenza A virus A/H3<br>Influenza A virus A/H1-2009<br>Influenza B virus<br>Parainfluenza virus 1<br>Parainfluenza virus 2<br>Parainfluenza virus 3<br>Parainfluenza virus 4<br>Respiratory syncytial virus<br>Human metapneumovirus |
| **DNA Virus** | | | Adenovirus |
| **Bacteria** | | | *Bordetella parapertussis*<br>*Bordetella pertussis*<br>*Chlamydia pneumoniae*<br>*Mycoplasma pneumoniae* |

**Table S2. Reference genomes used in the mapping pipeline.**

| Accession | Reference name | Routine PCR target | Length | Number of segments |
|---|---|---|---|---|
| MF502426.1 | Human_adenovirus_21_isolate_OHT_006_complete_genome | Adenovirus | 35341 | 1 |
| OR222614.1 | CP085971.1 | Bordetella parapertussis | 4775567 | 1 |
| OR222614.1 | NZ_CP025371.1 | Bordetella pertussis | 4088701 | 1 |
| BK015851.1 | NC_005043.1 | Chlamydia pneumoniae | 1225935 | 1 |
| NC_002645.1 | Human_coronavirus_229E | Coronavirus 229E | 27317 | 1 |
| NC_006577.2 | Human_coronavirus_HKU1 | Coronavirus HKU | 29926 | 1 |
| NC_005831.2 | Human_Coronavirus_NL63 | Coronavirus NL63 | 27553 | 1 |
| NC_006213.1 | Human_coronavirus_OC43_strain_ATCC_VR-759 | Coronavirus OC43 | 30741 | 1 |
| NC_002023.1;NC_002021.1;NC_002022.1;NC_002017.1;NC_002019.1;NC_002018.1;NC_002016.1;NC_002020.1 | Influenza A/H1 | Influenza A/H1 | 13588 | 8 |
| NC_026438.1;CY046941.1;NC_026437.1;NC_026433.1;NC_026436.1;CY121682.1;CY124074.1;GQ229379.2 | Influenza A/H1-2009 | Influenza A/H1-2009 | 13158 | 8 |
| NC_007373.1;NC_007372.1;NC_007371.1;NC_007366.1;NC_007369.1;NC_007368.1;NC_007367.1;NC_007370.1 | Influenza A/H3 | Influenza A/H3 | 13627 | 8 |
| NC_007357.1;NC_007358.1;NC_007359.1;NC_007362.1;NC_007360.1;NC_007361.1;NC_007363.1;NC_007364.1 | Influenza A/H5 | Influenza A/H5 | 13590 | 8 |
| NC_002205.1;NC_002206.1;NC_002207.1;NC_002208.1;NC_002209.1;NC_002210.1;NC_002211.1;NC_002204.1 | Influenza B | Influenza B | 14452 | 8 |
| NC_019843.3 | NC_019843.3 | MERS | 30119 | 1 |
| NC_039199.1 | Human_metapneumovirus_isolate_00-1 | Metapneumovirus | 13350 | 1 |
| BK056465.1 | NZ_LR214945.1 | Mycoplasma pneumoniae | 817125 | 1 |
| NC_003461.1 | Human_parainfluenza_virus_1 | Parainfluenza 1 | 15600 | 1 |
| NC_003443.1 | Human_rubulavirus_2 | Parainfluenza 2 | 15646 | 1 |
| NC_001796.2 | Human_parainfluenza_virus_3 | Parainfluenza 3 | 15462 | 1 |
| NC_021928.1 | Human_parainfluenza_virus_4a_viral_cRNA | Parainfluenza 4 | 17052 | 1 |
| U39662.1 | RSVA_Human_respiratory_syncytial_virus_S2 | RSV | 15190 | 1 |
| AY353550.1 | RSVB_Human_Respiratory_syncytial_virus_9320 | RSV | 15225 | 1 |
| NC_038306.1 | Human_coxsackievirus_A2_strain_Fleetwood | Rhinovirus/enterovirus | 7398 | 1 |
| NC_038308.1 | Human_enterovirus_68_strain_Fermon | Rhinovirus/enterovirus | 7367 | 1 |

| Accession | Reference name | Routine PCR target | Length | Number of segments |
|---|---|---|---|---|
| NC_030454.1 | Enterovirus_A114_strain_V13-0285 | Rhinovirus/enterovirus | 7317 | 1 |
| NC_001617.1 | Human_rhinovirus_89 | Rhinovirus/enterovirus | 7152 | 1 |
| KJ420749.1 | Enterovirus_sp._isolate_CPML_8109_08 | Rhinovirus/enterovirus | 7376 | 1 |
| NC_001430.1 | Human_enterovirus_D | Rhinovirus/enterovirus | 7390 | 1 |
| NC_001472.1 | Human_enterovirus_B | Rhinovirus/enterovirus | 7389 | 1 |
| NC_001612.1 | Human_enterovirus_A | Rhinovirus/enterovirus | 7413 | 1 |
| NC_038307.1 | Coxsackievirus_B3_mRNA | Rhinovirus/enterovirus | 7399 | 1 |
| NC_038312.1 | Human_rhinovirus_3 | Rhinovirus/enterovirus | 7208 | 1 |
| NC_009996.1 | Human_rhinovirus_C | Rhinovirus/enterovirus | 7099 | 1 |
| MW562722.1 | SARS-CoV-2 | SARS-CoV-2 | 29903 | 1 |
| NC_001417.2 | Phage MS2 genome | None – extraction and amplification control | 3569 | 1 |
| ATCC-VR-1838 | DQ Zika virus; Strain: MR 766 | None – amplification control | 10952 | 1 |
| ATCC-VR-824DQ | Reovirus Serotype 3; Strain: Dearing | None – amplification control | 23416 | 10 |
| ATCC-VR-907DQ | Sendai Virus | None - amplification control | 15362 | 1 |

**Table S3. Sensitivity and specificity of 8 different individual metrics to identify target pathogens in the derivation and validation sets.**

| | Derivation | | | | | Validation | | | | |
|---|---|---|---|---|---|---|---|---|---|---|
| Metric | Sensitivity N=67* | Sensitivity excluding true-positives with 0 reads N=49 | Specificity N=1154 | AUROC | Youden's optimal threshold | Sensitivity N=260 | Sensitivity excluding true positives with 0 reads N=171 | Specificity N=3910 | AUROC | Youden's optimal threshold |
| Area under the genome truncated at depth 10 | 0.716 | 0.878 | 0.991 | 0.856 | 0.003 | 0.642 | 0.807 | 0.981 | 0.816 | 0.004 |
| Percentage coverage at depth 1 | 0.716 | 0.878 | 0.991 | 0.856 | 0.25 | 0.642 | 0.807 | 0.981 | 0.816 | 0.4 |
| Number of reads mapping to the target reference as a percentage of the total number of reads in the run | **0.731** | 0.857 | 0.993 | **0.864** | 0.00004 | 0.658 | 0.795 | 0.978 | 0.823 | 0.00002 |
| Number of reads mapping to the target reference as a percentage of the number of reads mapping to any reference in the run | **0.731** | 0.878 | 0.993 | **0.864** | 0.006 | 0.658 | 0.895 | 0.978 | 0.824 | 0.001 |
| <u>Number of reads mapping to the target reference as a percentage of the number of reads mapping to the same reference in the run</u> | **0.731** | 0.980 | **0.995** | **0.864** | 0.037 | 0.658 | 0.936 | 0.979 | 0.822 | 0.003 |
| Number of reads mapping to the target reference as a percentage of the number of reads mapping to any reference in the same sample | **0.731** | **1.000** | 0.991 | 0.861 | 14.29 | 0.658 | 1.000 | 0.978 | 0.816 | 11.12 |
| Mean read length | 0.716 | 0.694 | 0.991 | 0.854 | 220 | 0.642 | 0.825 | 0.979 | 0.809 | 210 |
| Mean mapped read length | 0.716 | 0.878 | 0.991 | 0.855 | 70 | 0.642 | 0.848 | 0.982 | 0.812 | 90 |

* maximum possible 49/67=0.731 as 18 positives had 0 reads mapping to the target pathogen.

Note: AUROC=area under the receiver operating curve. Bold shows highest values for each individual metric in derivation set. See **Fig.S3** for the distribution of these metrics in the derivation set. Underline shows the alternative criterion used in an exploratory analysis of the validation set (**Tab.3**).

**Table S4. Sensitivity and specificity of 20 other individual metrics considered for target pathogen identification in the derivation set**

| | Derivation | | | | |
|---|---|---|---|---|---|
| Metric | Sensitivity N=67 | Sensitivity excluding true-positives with 0 reads N=49 | Specificity N=1154 | AUROC | Youden's optimal threshold |
| Mean non-zero mapped read depth across the whole genome | 0.716 | 0.796 | 0.991 | 0.856 | 1 |
| Mean non-zero mapped read depth across the whole genome truncated at depth 5 | 0.716 | 0.796 | 0.991 | 0.856 | 1 |
| Mean non-zero mapped read depth across the whole genome truncated at depth 10 | 0.716 | 0.796 | 0.991 | 0.856 | 1 |
| Area under the genome (not truncated) (including zero coverage regions) | 0.716 | 0.878 | 0.991 | 0.856 | 0.1 |
| Area under the genome truncated at depth 5 (including zero coverage regions) | 0.716 | 0.878 | 0.991 | 0.856 | 0.1 |
| Total number of mapped bases | 0.716 | 0.735 | 0.991 | 0.856 | 80 |
| Mapped bases as a percentage of reference length | 0.716 | 0.878 | 0.991 | 0.856 | 0.24 |
| Number of bases mapped at depth 1 | 0.716 | 0.735 | 0.991 | 0.856 | 71 |
| Number of bases mapped at depth 3 | 0.478 | 0.592 | 0.999 | 0.739 | 23 |
| Number of bases mapped at depth 5 | 0.373 | 0.510 | 1.000 | 0.687 | 246 |
| Number of bases mapped at depth 10 | 0.313 | 0.429 | 1.000 | 0.657 | 36 |
| Percentage coverage at depth 3 | 0.478 | 0.592 | 0.999 | 0.738 | 0.2 |
| Percentage coverage at depth 5 | 0.373 | 0.510 | 1.000 | 0.687 | 1.9 |
| Percentage coverage at depth 10 | 0.313 | 0.429 | 1.000 | 0.657 | 0.3 |
| Number of mapped reads | 0.731 | 0.755 | 0.991 | 0.864 | 1 |
| Number of mapped reads with alignment length ≥200 | 0.612 | 0.694 | 0.997 | 0.805 | 1 |
| Number of mapped reads with alignment length ≥300 | 0.522 | 0.633 | 0.999 | 0.761 | 1 |
| Number of mapped reads with alignment length ≥400 | 0.463 | 0.633 | 1.000 | 0.731 | 1 |
| Median length of sequenced reads | 0.716 | 0.735 | 0.991 | 0.853 | 209 |
| Median length of mapped read alignment length | 0.716 | 0.837 | 0.991 | 0.855 | 71 |

Note: AUROC=area under the receiver operating curve. See **Tab.S2** for results for eight prioritised metrics in derivation and validation sets and **Methods** for detailed definitions.

**Table S5. Sensitivity and specificity of combinations of 2-8 of the 8 prioritised metrics to identify target pathogens in the derivation set**

| Metric combination (see legend) | Sensitivity N=67 pathogen targets | Sensitivity excluding true-positives with 0 reads N=49 | Specificity N=1154 | AUROC |
|---|---|---|---|---|
| ('Sample_reads_percent_of_run', 'Sample_reads_percent_of_refs', 'Sample_reads_percent_of_type_run') | 0.731 | 0.959 | 0.993 | 0.862 |
| **('AuG_trunc10', 'Cov1_perc', 'Sample_reads_percent_of_refs')** | **0.731** | **0.898** | **0.991** | **0.861** |
| ('AuG_trunc10', 'Sample_reads_percent_of_run', 'Sample_reads_percent_of_refs') | 0.731 | 0.878 | 0.991 | 0.861 |
| ('AuG_trunc10', 'Sample_reads_percent_of_run', 'Sample_reads_percent_of_type_sample') | 0.731 | 1.000 | 0.991 | 0.861 |
| ('AuG_trunc10', 'Sample_reads_percent_of_run', 'mean_read_length') | 0.731 | 1.000 | 0.991 | 0.861 |
| ('AuG_trunc10', 'Sample_reads_percent_of_run', 'mean_aligned_length') | 0.731 | 0.898 | 0.991 | 0.861 |
| ('AuG_trunc10', 'Sample_reads_percent_of_refs', 'Sample_reads_percent_of_type_run') | 0.731 | 0.959 | 0.991 | 0.861 |
| ('AuG_trunc10', 'Sample_reads_percent_of_refs', 'Sample_reads_percent_of_type_sample') | 0.731 | 1.000 | 0.991 | 0.861 |
| ('AuG_trunc10', 'Sample_reads_percent_of_refs', 'mean_read_length') | 0.731 | 1.000 | 0.991 | 0.861 |
| ('AuG_trunc10', 'Sample_reads_percent_of_refs', 'mean_aligned_length') | 0.731 | 0.918 | 0.991 | 0.861 |
| ('AuG_trunc10', 'Sample_reads_percent_of_type_run', 'Sample_reads_percent_of_type_sample') | 0.731 | 1.000 | 0.991 | 0.861 |
| ('AuG_trunc10', 'Sample_reads_percent_of_type_run', 'mean_read_length') | 0.731 | 1.000 | 0.991 | 0.861 |
| ('AuG_trunc10', 'Sample_reads_percent_of_type_run', 'mean_aligned_length') | 0.731 | 0.959 | 0.991 | 0.861 |
| ('Cov1_perc', 'Sample_reads_percent_of_run', 'Sample_reads_percent_of_refs') | 0.731 | 0.898 | 0.991 | 0.861 |
| ('Cov1_perc', 'Sample_reads_percent_of_run', 'Sample_reads_percent_of_type_sample') | 0.731 | 1.000 | 0.991 | 0.861 |
| ('Cov1_perc', 'Sample_reads_percent_of_run', 'mean_read_length') | 0.731 | 1.000 | 0.991 | 0.861 |
| ('Cov1_perc', 'Sample_reads_percent_of_run', 'mean_aligned_length') | 0.731 | 0.918 | 0.991 | 0.861 |
| ('Cov1_perc', 'Sample_reads_percent_of_refs', 'Sample_reads_percent_of_type_run') | 0.731 | 0.980 | 0.991 | 0.861 |
| ('Cov1_perc', 'Sample_reads_percent_of_refs', 'Sample_reads_percent_of_type_sample') | 0.731 | 1.000 | 0.991 | 0.861 |
| ('Cov1_perc', 'Sample_reads_percent_of_refs', 'mean_read_length') | 0.731 | 1.000 | 0.991 | 0.861 |
| ('Cov1_perc', 'Sample_reads_percent_of_refs', 'mean_aligned_length') | 0.731 | 0.939 | 0.991 | 0.861 |
| ('Cov1_perc', 'Sample_reads_percent_of_type_run', 'Sample_reads_percent_of_type_sample') | 0.731 | 1.000 | 0.991 | 0.861 |
| ('Cov1_perc', 'Sample_reads_percent_of_type_run', 'mean_read_length') | 0.731 | 1.000 | 0.991 | 0.861 |
| ('Cov1_perc', 'Sample_reads_percent_of_type_run', 'mean_aligned_length') | 0.731 | 0.980 | 0.991 | 0.861 |
| ('Sample_reads_percent_of_run', 'Sample_reads_percent_of_refs', 'Sample_reads_percent_of_type_sample') | 0.731 | 1.000 | 0.991 | 0.861 |
| ('Sample_reads_percent_of_run', 'Sample_reads_percent_of_refs', 'mean_read_length') | 0.731 | 1.000 | 0.991 | 0.861 |
| ('Sample_reads_percent_of_run', 'Sample_reads_percent_of_refs', 'mean_aligned_length') | 0.731 | 0.918 | 0.991 | 0.861 |
| ('Sample_reads_percent_of_run', 'Sample_reads_percent_of_type_run', 'Sample_reads_percent_of_type_sample') | 0.731 | 1.000 | 0.991 | 0.861 |
| ('Sample_reads_percent_of_run', 'Sample_reads_percent_of_type_run', 'mean_read_length') | 0.731 | 1.000 | 0.991 | 0.861 |
| ('Sample_reads_percent_of_run', 'Sample_reads_percent_of_type_run', 'mean_aligned_length') | 0.731 | 0.959 | 0.991 | 0.861 |

Note: maximum AUROC of any single metric 0.864 (**Tab.S2**). Showing the first 30 combinations of 3 metrics from **Tab.S2** with AUROC >0.86. Bold shows main positivity criteria selected on the basis of the least collinearity across the three included components (**Tab.2**): alternative criteria is 'Sample_reads_percent_of_type_run' (Number of reads mapping to the target reference as a percentage of the number of reads mapping to the same reference in the run) alone (**Tab.3**).'Sample_reads_percent_of_run'= Number of reads mapping to the target reference as a percentage of the total number of reads in the run; 'Sample_reads_percent_of_refs'= Number of reads mapping to the target reference as a percentage of the number of reads mapping to any reference in the run; 'Sample_reads_percent_of_type_sample'= Number of reads mapping to the target reference as a percentage of the number of reads mapping to any reference in the same sample; 'AuG_trunc10'= Area under the genome truncated at depth 10; 'Cov1_perc'= Percentage coverage at depth 1; 'mean_read_length'= Mean read length; 'mean_aligned_length'=Mean mapped read length.

**Table S6. Assay costs.** Costs represent those at January 2026

| Item | Unit cost | Samples per unit | Cost per sample (£GBP) |
|---|---|---|---|
| Reagents and supplies | | | |
| MS2 Extraction Control | 621.76 | 100 | 6.21 |
| Viral control genomic RNA | 1631 | 1000 | 1.63 |
| M-SAN enzyme | 1,258.86 | 30 | 41.96 |
| Intact Virus Precipitation Reagent | 500.47 | 100 | 0.5 |
| Direct-zol(TM) RNA MicroPrep | 756 | 200 | 3.78 |
| Maxima H Minus First Strand cDNA Synthesis Kit | 637.52 | 100 | 6.37 |
| Sequenase Version 2.0 DNA Polymerase | 145.6 | 200 | 3.83 |
| AMPURE XP Magnetic Beads | 1220 | 400 | 3.05 |
| NEBNext(R) Ultra(TM) II Q5(R) Master Mix | 358.4 | 250 | 1.43 |
| Sub-total per sample | | | 68.77 |
| Adjustment by 20% to cover incidental costs, tips, power | | | 13.75 |
| Total reagents per sample | | | **82.52** |
| Sequencing consumables – assuming 32 samples per run | | | |
| ONT Rapid barcoding kit (SQK-RBK114.96) | 155.83 | | 4.33 |
| R10.4.1 Flow cell | 560 | | 15.56 |
| Total sequencing per sample | | | **19.89** |
| Infrastructure cost | | | |
| GridION purchase | 52,000 | | |
| Annual servicing | 5,600 | | |
| Total per sample (assuming 1 run per week and 5 year instrument life) | | | 9.61 |
| Total cost per sample (£GBP) | | | **112.02** |
| Assay time | | | |
| Nucleic acid for sequencing from sample (minutes per batch 16 samples) | | | |
| Host depletion and viral enrichment | 180 | | |
| RNA extraction | 90 | | |
| ss cDNA synthesis | 210 | | |
| ds cDNA synthesis | 180 | | |
| SISPA PCR, clean up and quantification | 330 | | |
| Per sample (minutes) | | | 61.9 |
| Long read sequencing library preparation (minutes per run 32 samples) | | | |
| DNA concentration normalisation | 90 | | |
| Sequencing library preparation | 180 | | |
| Per sample (minutes) | | | 8.44 |
| Total labour time per sample (minutes) | | | **70.3** |

**Figure S1. Total sequencing data yield and total number of reads per sample for derivation and validation sets.** Red dashed lines denote the thresholds set for a sequencing run to pass (minimum of 25k reads, and total yield of ≥400Mb, **Tab.2**). Square shapes represent batch negative controls and circle shapes represent sequences derived from nasopharyngeal samples.

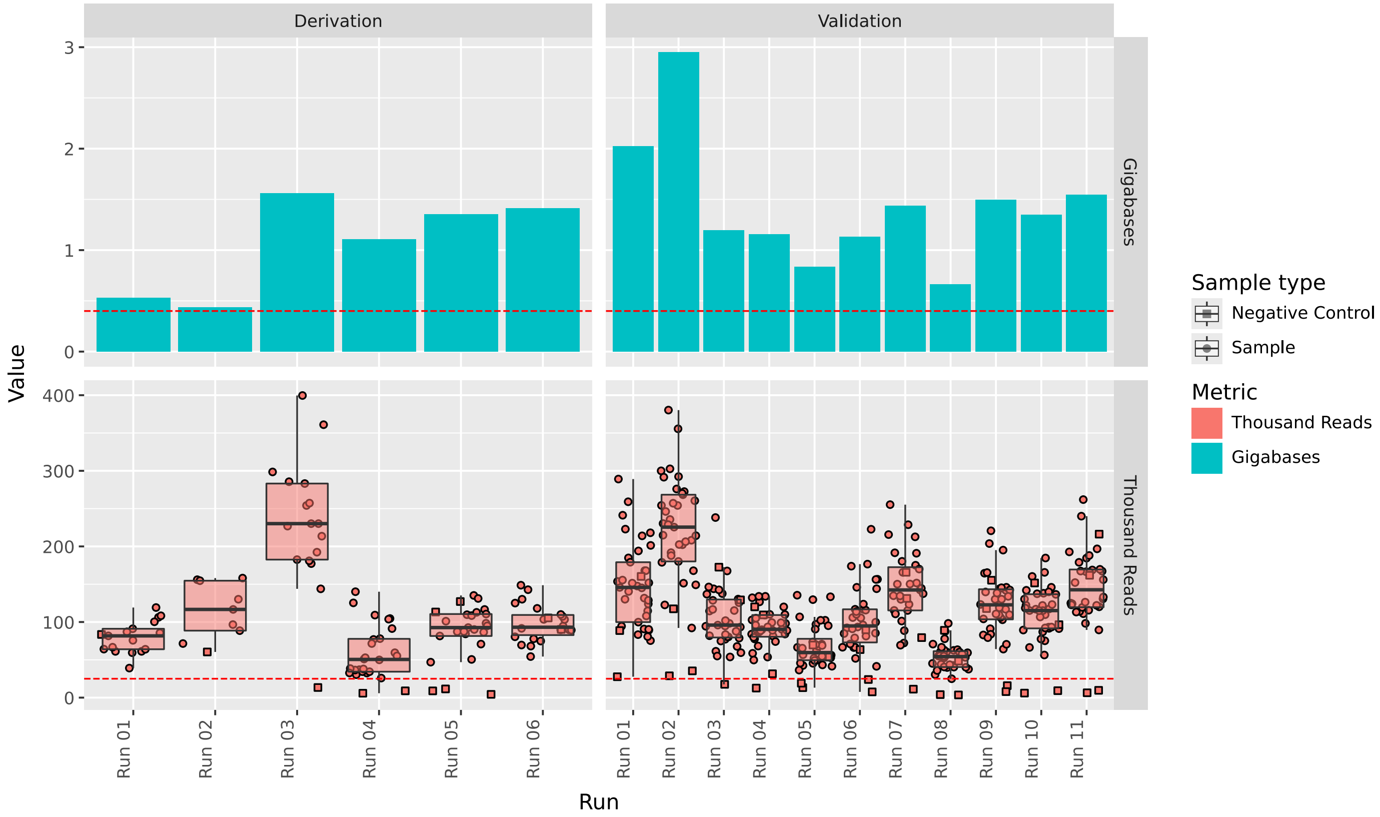

**Figure S2. Barcode cross-over in derivation set (before and after super accuracy base-calling) and in validation set (with super accuracy base-calling).**

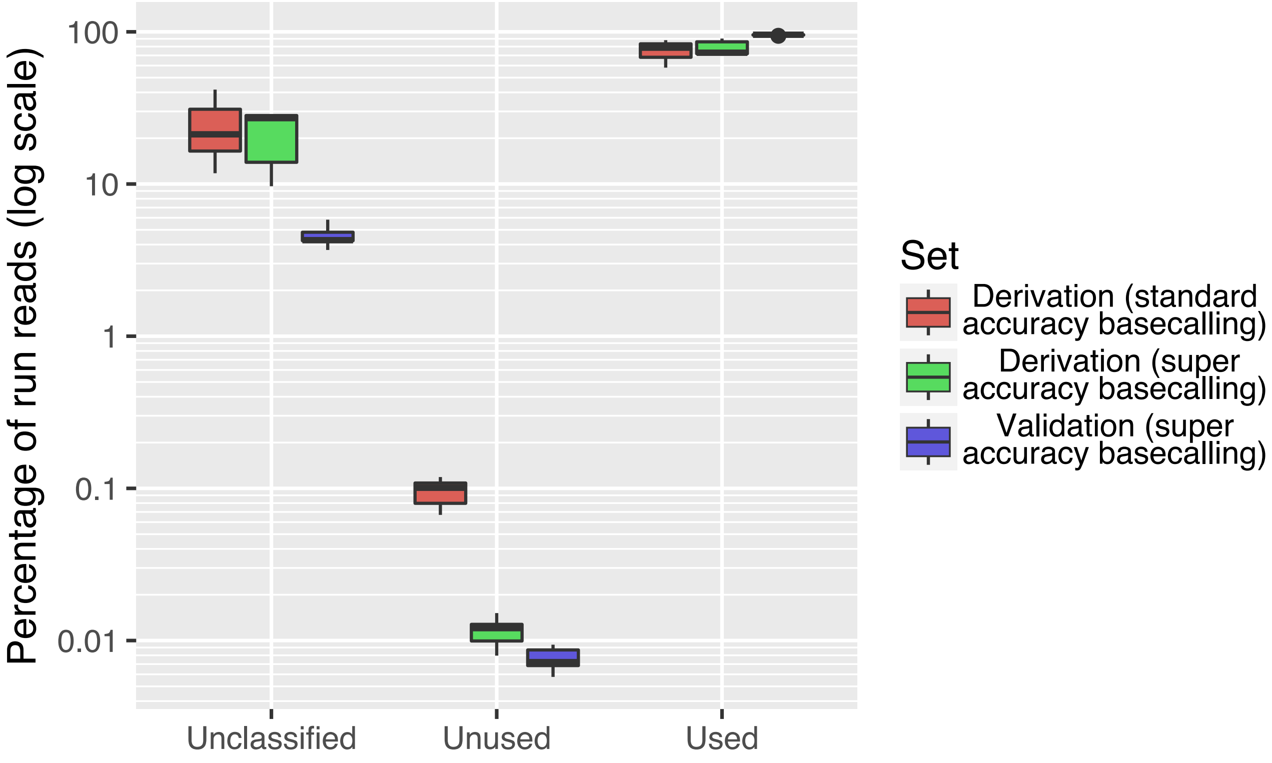

**Figure S3. Distribution of 8 key metrics in the derivation set.** Size of the circles represents magnitude of the Spearman correlation (r) between two metrics, symbols represent P value .<0.1, *<0.05, **<0.01, ***<0.001. Smoothed line by lowess using 0.5 fraction.

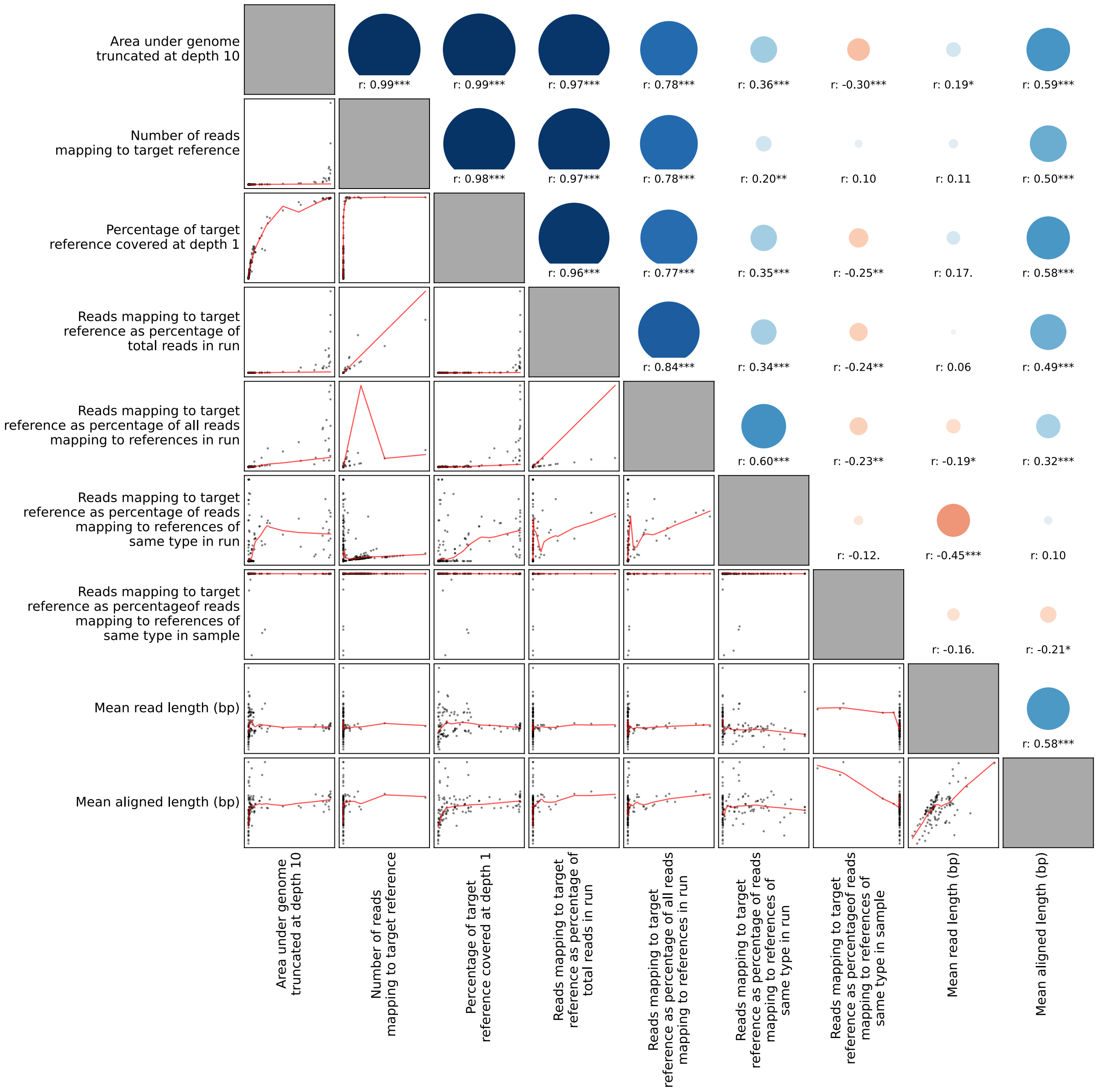

**Figure S4. Positivity detection using alternative criteria in the derivation set (A) and validation set (B).** Red line represents inclusion/exclusion threshold of 0.037%, derived from Youdens's index using the QC passed samples in the derivation set (**Tab.S2**). The sample QC pass contains samples that passed run, batch, and sample controls.

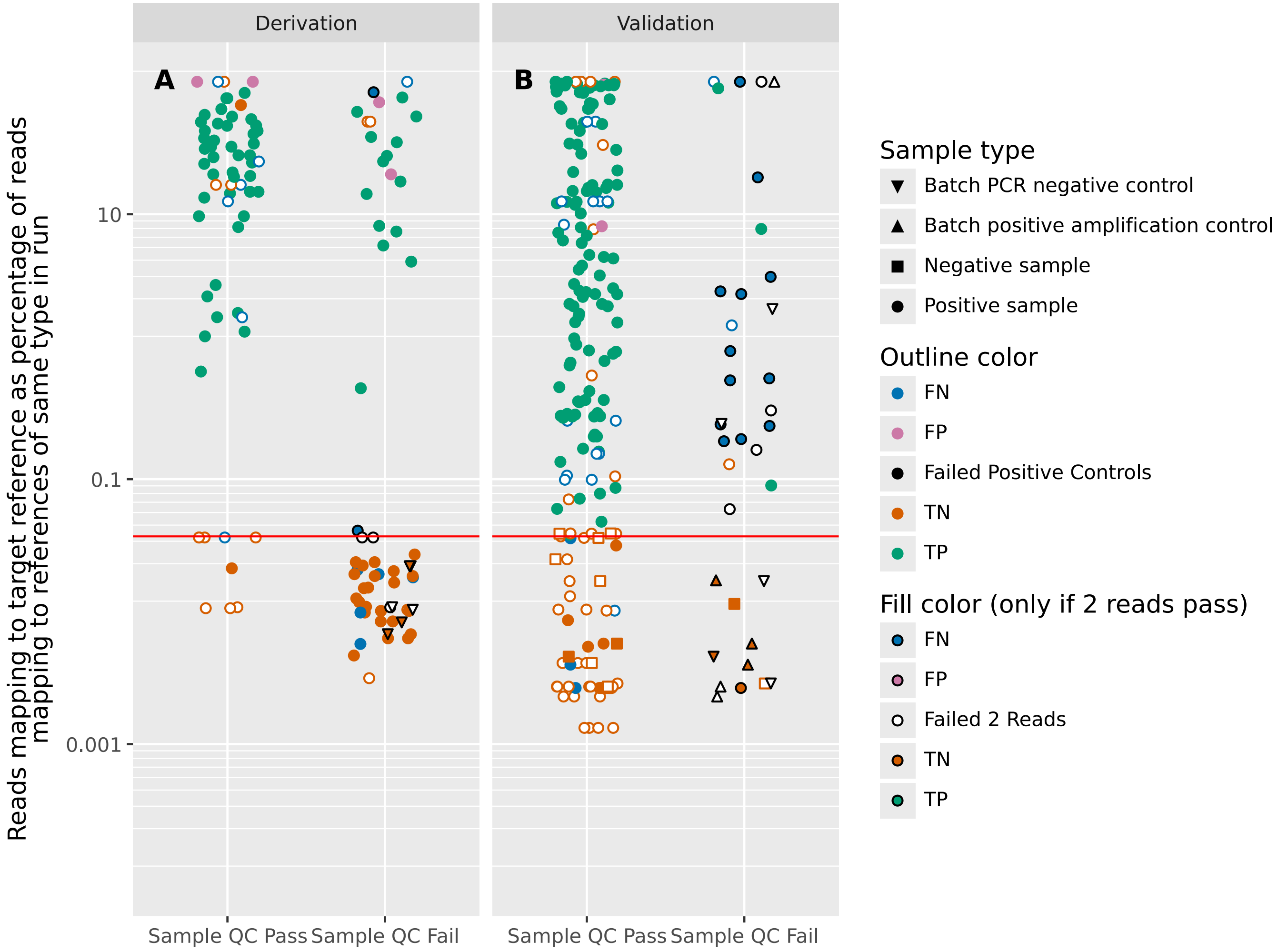

**Figure S5. Association between Cn/Ct values from routine diagnostic PCR assay (Resp-4-Plex/Xpert) and study qPCR (A), distribution of Cn/Ct values from both (B), and percentage of samples with target detected by study qPCR (C).** Spearman correlation in panel A 0.84, 0.55 and 0.91 for Influenza A, RSV and SARS-CoV-2 respectively.

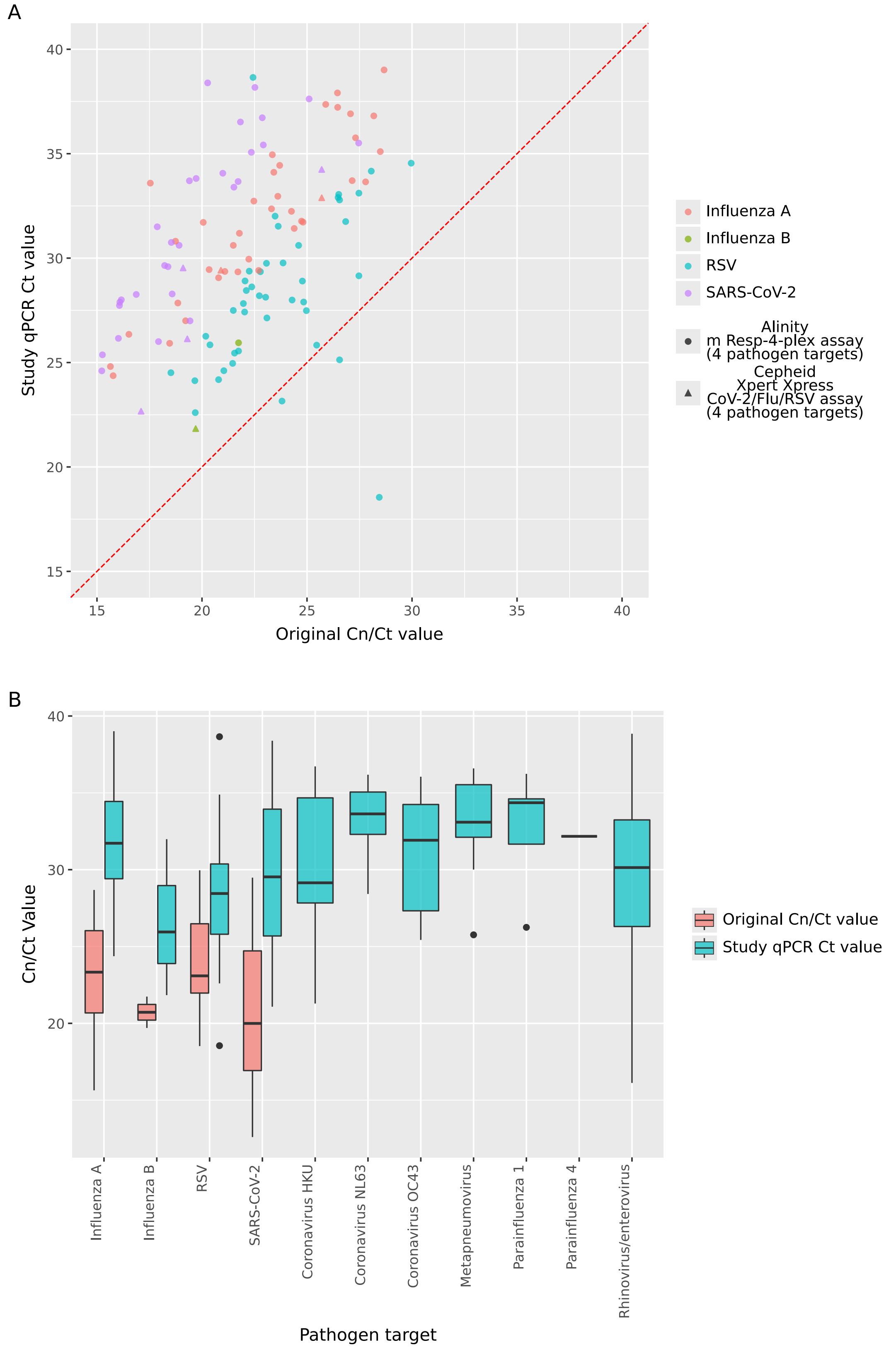

C

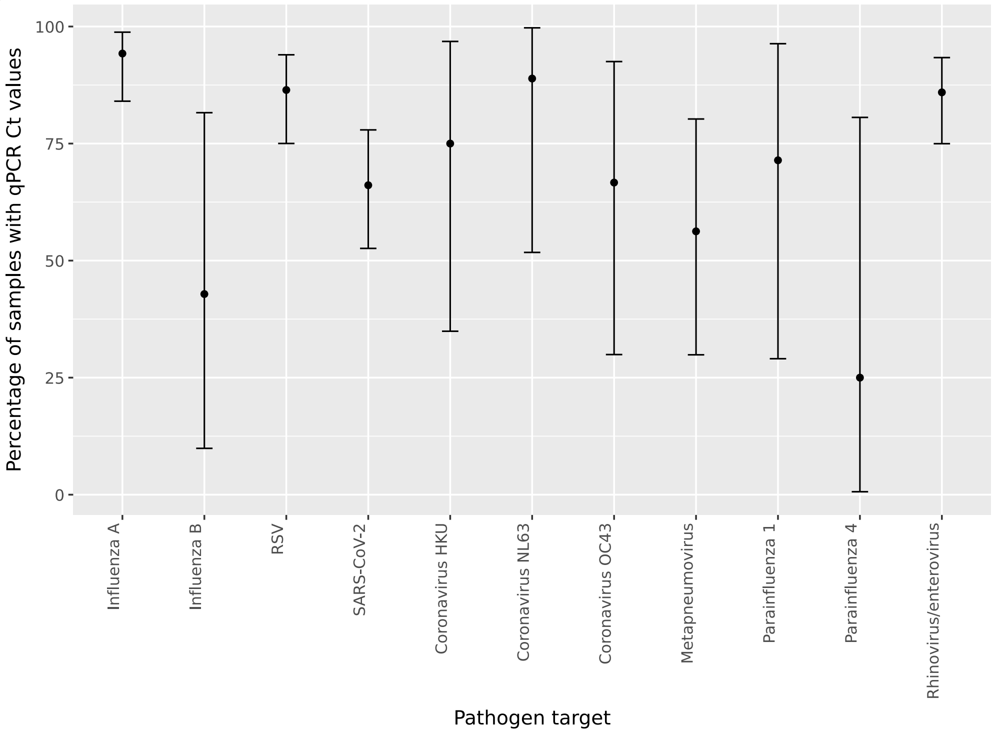
Percentage of samples with qPCR Ct values
100
75
50
25
0
Influenza A
Influenza B
RSV
SARS-CoV-2
Coronavirus HKU
Coronavirus NL63
Coronavirus OC43
Metapneumovirus
Parainfluenza 1
Parainfluenza 4
Rhinovirus/enterovirus
Pathogen target

**Figure S6. Mapping quality scores from aligned reads by the percentage difference to the reference genomes.** Dots represent reads mapping expectedly (species positive by gold standard, blue) and unexpectedly (species negative by gold standard, red). Red dashed line at mapQ=40 represents read mapping quality threshold applied to bacterial species only, to limit non-specific mapping.

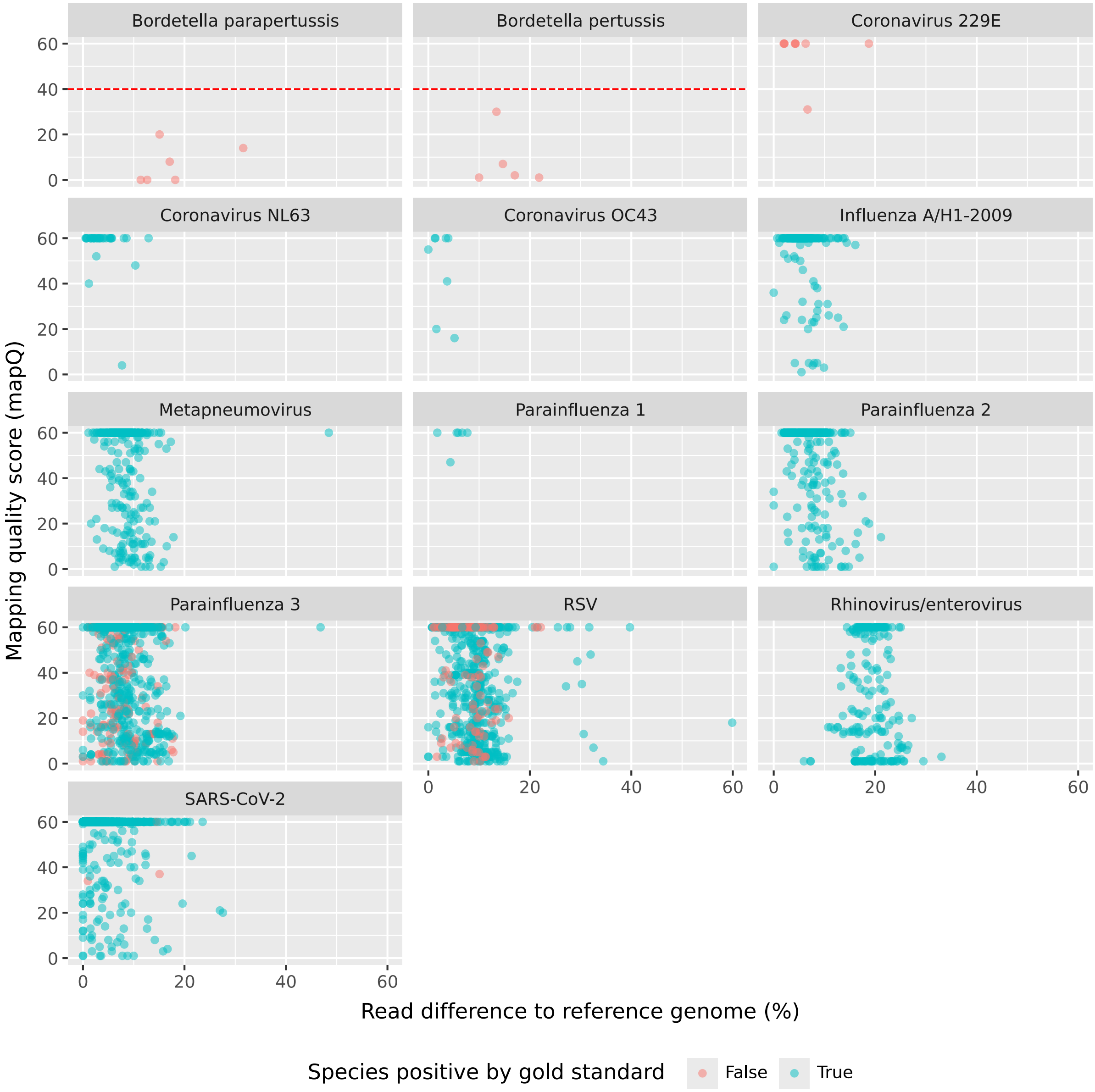
